# Risk factors for invasive pneumococcal disease in adults: a systematic review and meta-analysis

**DOI:** 10.1101/2025.03.13.25323815

**Authors:** Kim Ditzel, Federica Giardina, Jaap ten Oever, Amelieke J.H. Cremers

## Abstract

**Background:** The burden of invasive pneumococcal disease (IPD) in adults remains high despite vaccination programs. Age is currently used as a prime criterium for defining target groups for prevention. To support effective intervention programs, we studied the relative importance of risk conditions that influence susceptibility to adult IPD.

**Methods:** We conducted a systematic search in PubMed and Embase and included all original studies published before June 2024. We extracted the reported risk estimates for IPD in adults with risk conditions, compared to the general population (PROSPERO ID 417980). Meta-analyses were performed for risk conditions that were substantiated by more than one study, using pooled incidence rate ratios with 95% confidence intervals (IRRs (95%CIs)) as primary outcome.

**Findings:** Of the 2833 reports screened, 56 were included in the systematic review, and 45 supported the meta-analyses. The included articles reported more than 50 different risk factors for adult IPD. We synthesized 21 risk clusters for meta-analyses. The highest IRRs (95%CIs) for adult IPD were reported in immunocompromising conditions: transplant recipients 30·75 (17·64-53·60), asplenia 24·29 (18·63-31·65), HIV positive status 21·87 (15·72-30·43), and haematological malignancy 20·78 (9·94-43·47). Increasing age itself mediated minor risk sizes. At lower age the risk conditions conferred relatively higher risk ratios. Chronic kidney disease in adults <65 year old posed an IRR (95%CI) of 18·40 (11·38-29.74), compared to 5·12 (2·08-12·63) among those over 65. IRRs >10 were also observed for compromised cerebrospinal fluid barrier and Down’s syndrome. The overall quality of evidence was very low, mainly due to a high risk of bias and large between-study heterogeneity. Several studies indicated that patients with multimorbidity can accumulate risk for IPD.

**Interpretation:** This meta-analysis provides the relative importance of all reported risk factors for adult IPD. How risk conditions interact in cases of multimorbidity remains largely unknown.

**Funding:** Radboudumc Community for Infectious Diseases encouragement Grant.

**Research in context:** *Evidence before this study:* Current evidence lacks a systematic overview of risk factors for adult IPD that allows assessment of their relative importance. It is largely fragmented into single observational studies or reviews that focus on specific host qualities.

*Added value of this study:* Our comprehensive overview provides insight in the quantity, quality, and validity of evidence that supports risk conditions for adult IPD. In our meta-analyses we identified conditions that contribute to the risk of adult IPD, yet may ask for better appreciation. These include pronounced relative risk sizes in younger adults living with risk conditions, cumulative risks from multi-morbidity, and protective effects from healthy ageing.

*Implications of all the available evidence:* Populations that likely benefit from greater attention for prevention of IPD are the younger adults living with risk conditions - especially chronic kidney disease, homeless individuals, and those with Down’s syndrome. In combination with absolute risk sizes, the provided risk ratio’s indicate the adult populations most vulnerable to IPD. In addition, risk sizes inform study designs concerning effectiveness of preventive strategies.

## Introduction

The bacterium *Streptococcus pneumoniae* (the pneumococcus) remains the major cause of respiratory sepsis and its related mortality.^1,2^ Each year, pneumococcal infections are responsible for an estimated 8·1 million disability-adjusted life-years, 829,000 deaths, and 40·3 million life years lost globally.^3,4^ Also in bacterial meningitis, the highest proportion of deaths remains attributable to *S. pneumoniae*.^5^ Invasive pneumococcal disease (IPD) represents a subset of pneumococcal infections in which the pneumococcus is identified in a human specimen collected from a normally sterile body site, using laboratory methods. This case definition ascertains the pneumococcal aetiology of a given infection and delineates a population for epidemiology purposes.

Individuals at the youngest and oldest age groups bear the largest IPD burden. In 2000 the first conjugated pneumococcal vaccine (PCV) was introduced into a paediatric national immunization programme with primary objective to protect children below the age of five from IPD.^6^ In this age category high efficacy was demonstrated for protection from IPD caused by serotypes included in the heptavalent vaccine.^7^ Serotype-based vaccination not only altered the pneumococcal population that infected the target group itself, but similarly changed the population infecting adults living in the same area.^8^ This phenomenon is called “herd protection” and it suggests that the paediatric nasopharyngeal niche is a relevant reservoir for pneumococcal acquisition in adults.^9^ However, what differentiated adults from the paediatric target group, was the more comprehensive replacement of the adult IPD case load by non-vaccines serotypes in many areas in the world.^10,11^ The largely sustained burden of IPD among adults substantiates the potential value of preventive measures that target this specific age category.

The replacing pneumococcal population in adult IPD is characterized by a diversification of pneumococcal serotypes.^12,13^ This relative lack of dominant serotypes not only complicates the design and coverage of next generation vaccines that target adults.^14,15^ It also suggests that in adults, the permissiveness of the human host to infection could be a stronger determinant of IPD compared to children where the range of potentially pathogenic pneumococcal serotypes seems notably more confined thus far.

Over the past years several novel pneumococcal vaccines with extended serotype valency were submitted for licensure to the medical regulatory authorities. In adults, age is currently the primary criterion used to define target groups for interventions aimed at preventing adult IPD. However, if additional risk conditions significantly contribute to adults’ susceptibility to IPD, risk prediction could be improved. In addition, the effectiveness of preventive interventions may depend on the specific vulnerability that mediates the risk of IPD. Currently, 54 countries have established indications for adult pneumococcal vaccination in their national immunisation schedule, 38 of which have specified adult risk groups.^16^ In practice, these specified target groups for prevention of adult IPD are regularly derived from evidence generated for more narrow study populations and associated risk sizes are rarely accounted for. Optimizing the effectiveness of interventions requires the identification and prioritization of adults who carry marked risks for IPD. However, current evidence on potential risk factors for adult IPD is largely fragmented and does not provide insight in risk sizes across risk conditions.

Therefore, the aim of this systematic review and meta-analyses of observational studies was to provide a comprehensive overview of all reported risk conditions for adult IPD, and to compare their relative risk size.

## Methods

### Search strategy and selection criteria

This systematic review and the meta-analyses of observational studies were all conducted in accordance with the COSMOS-E guidelines and reported as advised by the PRISMA 2020 statement.^17,18^

Our study protocol was registered in PROSPERO on April 28^th^ 2023 (ID 417980). The search strategy and eligibility criteria for inclusion in the systematic review are detailed in Supplementary material A. In short, in April 2023 PubMed and Embase were searched for articles published in English language using extensive search terms for IPD in combination with risk factors, comorbidities and immunodeficiencies. To ensure the search strategy was sensitive enough to capture all key studies, it was iteratively refined in collaboration with the library at Radboudumc. In addition, the bibliographies of all relevant review articles were screened for missing, relevant articles. We did not search for grey literature (e.g. conference abstracts, preprints). Trial registries were not searched as the current study on risk conditions relied on cohort studies and case-control studies. An updated search was performed on June 11^th^ 2024.

All articles published up to the search date were assessed for eligibility using clear criteria (Supplementary material A). One of two researchers (K.D. and A.C.) performed a first round of unambiguous exclusions based on screening of titles and abstracts. In case of any doubt on potential eligibility, early exclusion was discussed for consensus between the two researchers. In brief, articles were potentially eligible if they contained primary data that provided summary estimates of a risk factor for adult IPD (not including non-invasive pneumococcal pneumonia) in comparison to the general population. The reason to exclude studies that report on host polymorphisms as a risk condition for IPD is that detailed systematic reviews already emphasized limitations of the existing evidence which is being addressed in a prospective manner.^19,20^

We excluded studies with ≥10% of the population aged ≤14 years. This represents a post-hoc amendment to the registered PROSPERO protocol, which initially stated a 18-year cutoff. This adjustment prevented the exclusion of some highly relevant studies.

A detailed assessment of eligibility was performed on full-text articles, and studies were included if independently approved by both investigators. If risk factors were supported by single studies or if determinants were defined too heterogeneously, the results were reported separately and excluded from the meta-analysis.

### Data analysis

Data were extracted by one of two researchers (K.D. or A.C.) and cross-checked by the other using a standardised form. The outcome variable was the risk of IPD, expressed as an incidence rate ratio (IRR), risk ratio (RR), hazard ratio (HR) or odds ratio (OR) with their corresponding 95% confidence interval, associated with a given risk condition. Both adjusted and unadjusted effect estimates were extracted, along with those reported for specific subpopulations (e.g., age categories). Additionally extracted data included study design, study population characteristics (e.g., period, country, control population, reported pneumococcal vaccination policy), definition of IPD, number of IPD patients, and sponsoring by commercial entities.

Given the substantial number of identified risk factors, we grouped them into broader categories – referred to as “risk clusters” – based on consensus. For studies with overlapping population and time periods, we selected only one study using the following hierarchical criteria: largest sample size, highest number of risk factors, most recent study, and availability of age-stratified data. Studies reporting on a combination of risk conditions spanning multiple risk clusters (e.g., haematological and solid malignancy combined) were excluded. For studies reporting risk estimates across multiple time periods, we prioritized the overall estimate when available. If an overall estimate was not provided, we selected the estimate for the most recent period, as we considered it the most representative.

The risk of bias in included studies was assessed using the Risk Of Bias In Non-randomised Studies - of Exposure (ROBINS-E) tool.^21^ This tool considers seven domains with a potential for bias: confounding, measurement of the exposure, selection of participants, post-exposure interventions, missing data, measurement of the outcome and selection of the reported result. Two researchers (K.D. and A.C.) independently evaluated the individual studies, resolved any differences at the domain level, and consolidated the risk of bias in the study to be low, of some concern, high, or very high according to the ROBINS-E tool.

The primary outcomes of interest (pooled IRR or RR) were stratified by age group, where applicable, into ‘all ages’, ‘age ≥65’ and ‘age <65’. For studies reporting ORs, these were approximated as RRs under the assumption of rare outcomes (incidence ≤5%).

For studies reporting risk estimates disaggregated into more refined categories than our predefined groups (e.g., age <65, ≥65, or all ages), we combined these estimates to align with our unit of analysis. First, the standard error (SE) of each risk estimate (e.g., RR or IRR) was derived from its reported confidence interval (CI) by transforming to the logarithmic scale and assuming a Wald confidence interval. Subsequently, multiple estimates within the same group were combined using standard inverse-variance weighting, under the assumption of independence. This approach utilized weighted least squares to compute a single combined estimate. Calculations were performed using the *escalc* and *aggregate* functions in the metafor package in R (version 4.4.0).

Random-effects models were used to pool effect estimates, accounting for between-study heterogeneity. Pooled estimates were calculated separately for IRRs and RRs, where reported. Heterogeneity was assessed using the I^2^ statistic, with thresholds interpreted as low (25%), moderate (50%), or high (75%) heterogeneity. Forest plots were generated to visualize pooled estimates. All analyses were performed using the metafor package.

For each risk cluster, the certainty of the body of evidence provided by the meta-analyses on IRRs was assessed using the Grades of Recommendation, Assessment, Development, and Evaluation (GRADE) system.^22^ Imprecision was defined as serious if the one-sided confidence interval was >45% of the pooled IRR for all ages, and as very serious if >65%.

## Results

Our search identified 2833 unique records (Figure 1). After screening titles and abstracts, we assessed full-text reports of 234 studies for eligibility. Two additional studies were identified through screening of citations. In total, 62 eligible publications were identified, representing 56 unique study populations, of which 45 were included in the meta-analyses. Studies excluded during full-text assessment, along with the reasons for exclusion, are detailed in Supplementary material B.

**Figure 1:**
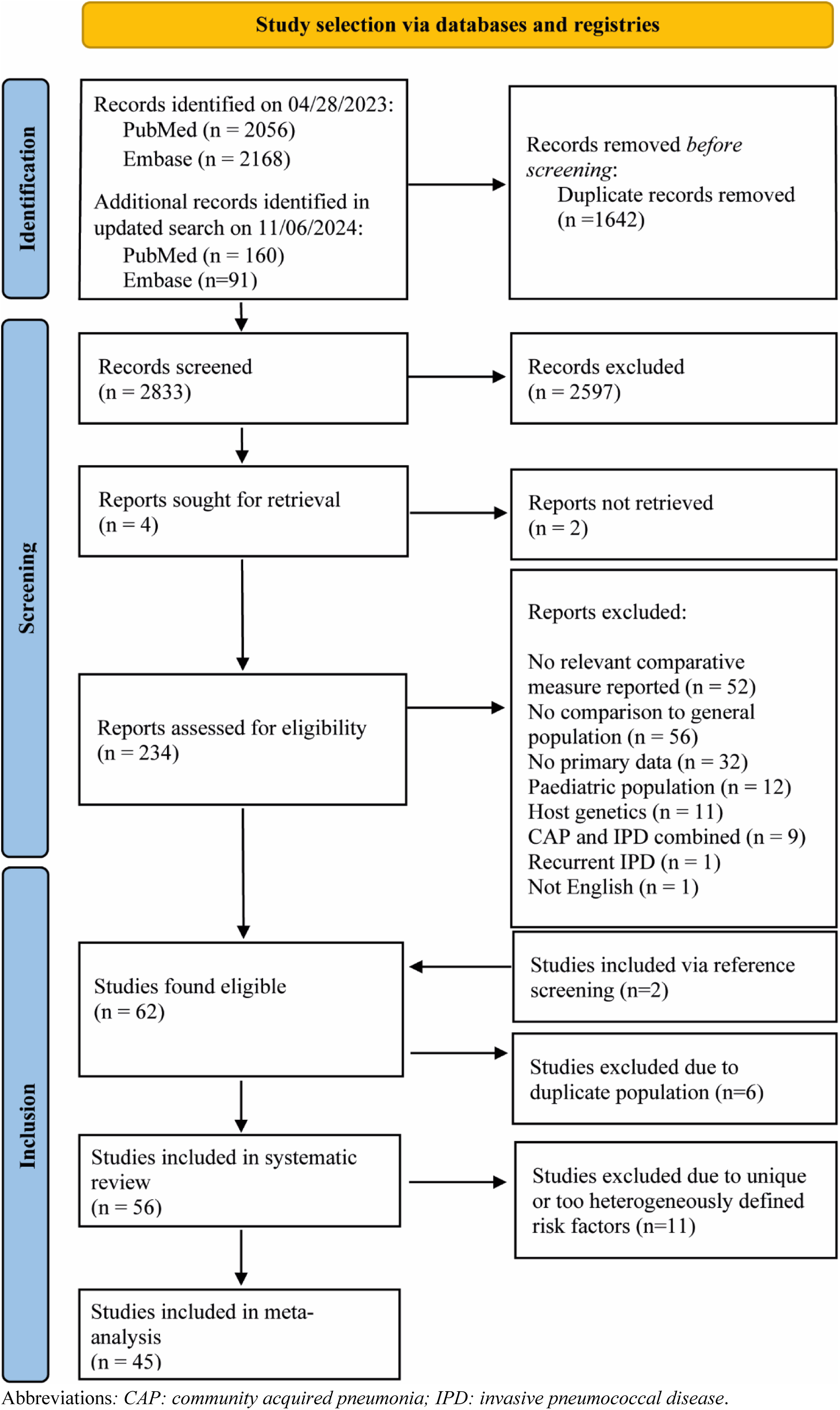
Study selection. The flow chart demonstrates the process by which studies were selected for the systematic review and for the meta-analyses.

The 56 studies included in the systematic review comprised cohort studies or case-control studies (Table 1). All were performed in high- or upper-middle income countries (Supplementary material C). The majority of studies was conducted more than 20 years ago, during periods with varying pneumococcal vaccination policies. IPD was uniformly defined as a positive culture with *S. pneumoniae* from a normally sterile site, for which seven studies relied on corresponding ICD-codes. Control groups ranged from matched controls to healthcare repositories or national surveillance data. Most studies investigated multiple potential risk factors and 39% of studies stratified risk factors by age. One third of the studies (17/56) was funded by commercial entities.

**Table 1:** Studies included in the systematic review.

| Study | Country | Study design | Years | IPD definition | n/N | Number of reported risk factors<sup>#</sup> |  |  |  |  |  |
| --- | --- | --- | --- | --- | --- | --- | --- | --- | --- | --- | --- |
|  |  |  |  |  |  | I | II | III | IV | V | VI |
| Steinberg et al, 2024<sup>23</sup> | United States | Cohort | 2011-2020 | Positive cultures | 1164/u | 0 | 0 | 0 | 0 | 0 | 2 |
| Grant et al, 2023<sup>24</sup> | United States | Cohort | 2016-2019 | ICD-codes | u/38618534* | 0 | 4 | 5 | 2 | 2 | 2 |
| Torén et al (a), 2022<sup>25</sup> | Sweden | Case control | 2006-2019 | Positive cultures | 3184/19243 | 0 | 0 | 0 | 5 | 0 | 0 |
| Torén et al (b), 2022<sup>26</sup> | Sweden | Case control | 2006-2019 | Positive cultures | 3968/21666 | 0 | 0 | 0 | 0 | 0 | 9 |
| Torén et al (c), 2022<sup>27</sup> | Sweden | Case control | 2007-2018 | Positive cultures | 6870/48090 | 0 | 3 | 4 | 1 | 0 | 7 |
| Deb et al, 2022<sup>28</sup> | Germany | Cohort | 2016-2019 | ICD-codes | 1053/3858335 | 0 | 0 | 0 | 0 | 0 | 2 |
| Kobayashi et al, 2022<sup>29</sup> | United States | Cohort | 2008-2018 | Positive cultures | 29668/u | 0 | 1 | 0 | 0 | 0 | 0 |
| Garcia Garrido et al, 2021<sup>30</sup> | Netherlands | Cohort | 2004-2016 | Positive cultures | 7167/39183860* | 0 | 2 | 0 | 0 | 0 | 0 |
| Kirwan et al, 2021<sup>31</sup> | United Kingdom | Cohort | 1999-2017 | Positive cultures | 108223/u | 0 | 1 | 0 | 0 | 0 | 0 |
| Winje et al, 2021<sup>32</sup> | Norway | Cohort | 2015-2017 | Positive cultures | 1020/u | 1 | 3 | 5 | 1 | 0 | 4 |
| Andersen et al, 2020<sup>33</sup> | Denmark | Cohort | 2000-2016 | Positive cultures | 13332/85002224* | 0 | 2 | 0 | 0 | 0 | 0 |
| Torén et al (d), 2020<sup>34</sup> | Sweden | Case control | 2006-2014 | Positive cultures | 4438/29956 | 0 | 0 | 0 | 1 | 0 | 0 |
| Lin et al, 2019<sup>35</sup> | Taiwan | Case control | 2000-2013 | ICD-codes | u/2265 | 0 | 1 | 4 | 0 | 0 | 1 |
| Dirmesropian et al, 2019<sup>36</sup> | Australia | Cohort | 2001-2015 | Positive cultures | 157/1.85 million | 0 | 0 | 0 | 0 | 0 | 1 |
| Zhang et al, 2018<sup>37</sup> | United States | Cohort | 2012-2014 | ICD codes | u/57108316* | 0 | 3 | 6 | 0 | 0 | 5 |
| Wiese et al, 2018<sup>38</sup> | United States | Case control | 1995-2014 | Positive cultures | 1233/25632 | 0 | 0 | 0 | 0 | 0 | 1 |
| Imai et al, 2018<sup>39</sup> | Japan | Cohort | 2006-2014 | ICD-codes | 197/9.3 million | 0 | 0 | 5 | 0 | 0 | 5 |
| Röckert Tjernberg et al, 2017<sup>40</sup> | Sweden | Case control | 1969-2009 | Positive cultures | 207/173269 | 0 | 0 | 0 | 0 | 0 | 1 |
| Backhaus et al, 2016<sup>41</sup> | Sweden | Cohort | 1996-2008 | Positive cultures | 2977/1512233 | 0 | 2 | 0 | 0 | 0 | 0 |
| Baxter et al, 2016<sup>42</sup> | United States | Cohort | 2008-2014 | Positive cultures | 1549/15102047* | 4 | 10 | 10 | 2 | 1 | 7 |
| Meiring et al, 2016<sup>43</sup> | South Africa | Cohort | 2003-2008 | Positive cultures | 4734/u | 0 | 1 | 0 | 0 | 0 | 0 |
| Wagenvoort et al, 2016<sup>44</sup> | Netherlands | Cohort | 2008-2012 | Positive cultures | 2224/174274 | 0 | 2 | 4 | 0 | 0 | 1 |
| Weycker et al, 2016<sup>45</sup> | United States | Cohort | 2007-2010 | ICD- codes | u/91.6 million* | 0 | 0 | 0 | 0 | 1 | 1 |
| Shigayeva et al, 2016<sup>46</sup> | Canada | Cohort | 1995-2012 | Positive cultures | 7902/4085487 | 0 | 7 | 1 | 0 | 0 | 0 |
| De st Maurice et al, 2016<sup>47</sup> | United States | Cohort | 1998-2013 | Positive cultures | 8383/2.5 million | 1 | 0 | 0 | 0 | 0 | 0 |
| Kantsø et al, 2015<sup>48</sup> | Denmark | Case control | 1977-2013 | Positive cultures | 4261/1556519 | 0 | 2 | 0 | 0 | 0 | 0 |
| Kwak et al, 2014<sup>49</sup> | South Korea | Cohort | 2010-2011 | ICD- codes | 826/1887401 | 0 | 0 | 2 | 0 | 0 | 0 |
| Harboe et al, 2014<sup>50</sup> | Denmark | Cohort | 1995-2010 | Positive cultures | 273/1112653* | 0 | 1 | 0 | 0 | 0 | 0 |
| Shea et al, 2014<sup>51</sup> | United States | Cohort | 2007-2010 | ICD- codes | u/91.6* | 0 | 7 | 7 | 2 | 1 | 5 |
| Inghammar et al, 2013<sup>52</sup> | Sweden | Case control | 1990-2008 | Positive cultures | 4085/44438 | 0 | 7 | 5 | 1 | 0 | 0 |
| Feemster et al, 2013<sup>53</sup> | United states | Cohort | 2005-2008 | Positive cultures | 1416/3849071 | 0 | 0 | 0 | 0 | 0 | 4 |
| Chapman et al, 2013<sup>54</sup> | United Kingdom | Cohort | 2006-2011 | Positive cultures | 1351/2148629 | 0 | 0 | 0 | 0 | 0 | 3 |
| van Hoek et al, 2012<sup>55</sup> | England | Cohort | 2002-2009 | Positive cultures | 20791/42105600 | 0 | 2 | 5 | 0 | 0 | 1 |
| Jacups et al, 2011<sup>56</sup> | Australia | Cohort | 1987-2008 | Positive cultures | 205/114537 | 2 | 0 | 1 | 2 | 0 | 1 |
| Klemets et al (a), 2010<sup>57</sup> | Finland | Case control | 1995-2002 | Positive cultures | 1282/14067 | 0 | 5 | 4 | 0 | 0 | 0 |
| Wong et al (a), 2010<sup>58</sup> | Canada | Cohort | 2000-2004 | Positive cultures | 863/u | 0 | 0 | 0 | 1 | 0 | 3 |
| Wong et al (b), 2010<sup>59</sup> | Canada | Cohort | 2000-2004 | Positive cultures | 1273/3 million | 0 | 2 | 0 | 0 | 0 | 0 |
| Metlay et al, 2010<sup>60</sup> | United States | Cohort | 2005-2008 | Positive cultures | 1376/3.7 million | 3 | 0 | 2 | 1 | 0 | 2 |
| Flory et al, 2008<sup>61</sup> | United States | Cohort | 2002-2004 | Positive cultures | 609/2881132 | 2 | 0 | 2 | 1 | 0 | 5 |
| Kumar et al (b), 2008<sup>62</sup> | Canada | Cohort | 1995-2004 | Positive cultures | 4073/5 million | 0 | 1 | 0 | 0 | 0 | 0 |
| Klemets et al (b), 2008<sup>63</sup> | Finland | Cohort | 1995-2002 | Positive cultures | 3624/5.2 million | 1 | 0 | 0 | 0 | 0 | 0 |
| Kumar et al (a), 2007<sup>64</sup> | Canada | Cohort | 1995-2004 | Positive cultures | 4073/4 million | 0 | 1 | 0 | 0 | 0 | 0 |
| Watt et al, 2007<sup>65</sup> | United States | Case control | 1999-2002 | Positive cultures | 118/471 | 0 | 0 | 9 | 4 | 0 | 9 |
| Talbot et al, 2005<sup>66</sup> | United States | Case control | 1995-2002 | Positive cultures | 451/4961 | 0 | 0 | 1 | 0 | 0 | 0 |
| Kyaw et al, 2005<sup>67</sup> | United States | Cohort | 1999-2000 | Positive cultures | 4335/14.9 million | 2 | 3 | 3 | 1 | 0 | 4 |
| Thomsen et al, 2004<sup>68</sup> | Denmark | Case control | 1992-2001 | Positive cultures | 598/6578 | 0 | 0 | 1 | 0 | 0 | 0 |
| Flannery et al, 2004<sup>69</sup> | United States | Cohort | 1998-2002 | Positive cultures | 15923/16.5 million | 1 | 0 | 0 | 0 | 0 | 0 |
| Kupronis et al, 2003<sup>70</sup> | United States | Cohort | 2000-2001 | Positive cultures | 1258/2.4 million | 0 | 0 | 0 | 0 | 0 | 1 |
| Nuorti et al (a), 2000<sup>71</sup> | United States | Cohort | 1994-1997 | Positive cultures | 399/735315 | 2 | 1 | 0 | 0 | 0 | 0 |
| Nuorti et al (b), 2000<sup>72</sup> | Canada and<br>United States | Case control | 1995-1996 | Positive cultures | 228/529 | 1 | 0 | 0 | 3 | 0 | 3 |
| Harrison et al, 2000<sup>73</sup> | United States | Cohort | 1995-1996 | Positive cultures | 1051/5 million | 1 | 0 | 0 | 0 | 0 | 1 |
| Sankilampi et al, 1997<sup>74</sup> | Finland | Cohort | 1983-1992 | Positive cultures | 934/1146843 | 1 | 0 | 0 | 0 | 0 | 0 |
| Plouffe et al, 1996<sup>75</sup> | United states | Cohort | 1991-1994 | Positive cultures | 419/u | 2 | 1 | 0 | 0 | 0 | 0 |
| Torzillo et al, 1995<sup>76</sup> | Australia | Cohort | 1985-1991 | Positive cultures | 78/31510 | 0 | 0 | 0 | 0 | 0 | 1 |
| Haglund et al, 1993<sup>77</sup> | United States | Cohort | 1989-1990 | Positive cultures | 144/848273 | 0 | 0 | 0 | 0 | 0 | 1 |
| Bennett et al, 1992<sup>78</sup> | United States | Cohort | 1985-1989 | Positive cultures | 427/712695 | 1 | 0 | 0 | 0 | 0 | 0 |

Positive cultures were defined as positive cultures from a normally sterile body site.
n: number of IPD cases; N: total number of participants; u: unknown; \*: person-years.

The risk of bias of the included articles ranged from “high” to “very high” based the on ROBINS-E assessment tool. The major sources of bias were an observational study nature, the size of potential confounding, and unknown allocation of post-exposure interventions such as vaccination, prophylactic antibiotics and treatment of the underlying disease. Even though 77% (43/56) of the studies attempted to adjust for confounders like age, in all of those studies the risk of bias was still assessed as “high” (25/43) to “very high” (18/43). A summary of the risk of bias assessment is represented in supplementary material D.

The 56 studies reported on >50 potential risk conditions for adult IPD, plus further subgroups. We binned risk conditions into 21 risk clusters for which meta-analyses were performed. As factor age affected the risk of adult IPD (Supplementary material E) and because many included studies reported risk estimates stratified by age, we followed that approach. From the 45 studies included in the meta-analyses, 212 eligible incidence rate ratios (IRRs) and 111 odds ratios (ORs) were extracted (complete data provided in Supplementary material F). The number of risk estimates extracted per risk cluster varied between 2 and 38 (Figure 2). Serious concern on the occurrence of a risk condition in the control population was present in one case-control study, that relied on discharge diagnoses assigned over a short 12-months period for three low prevalent risk conditions – which risk estimates were therefore removed from the meta-analyses. Otherwise, no other risk estimates were excluded from the meta-analyses.

**Figure 2:**
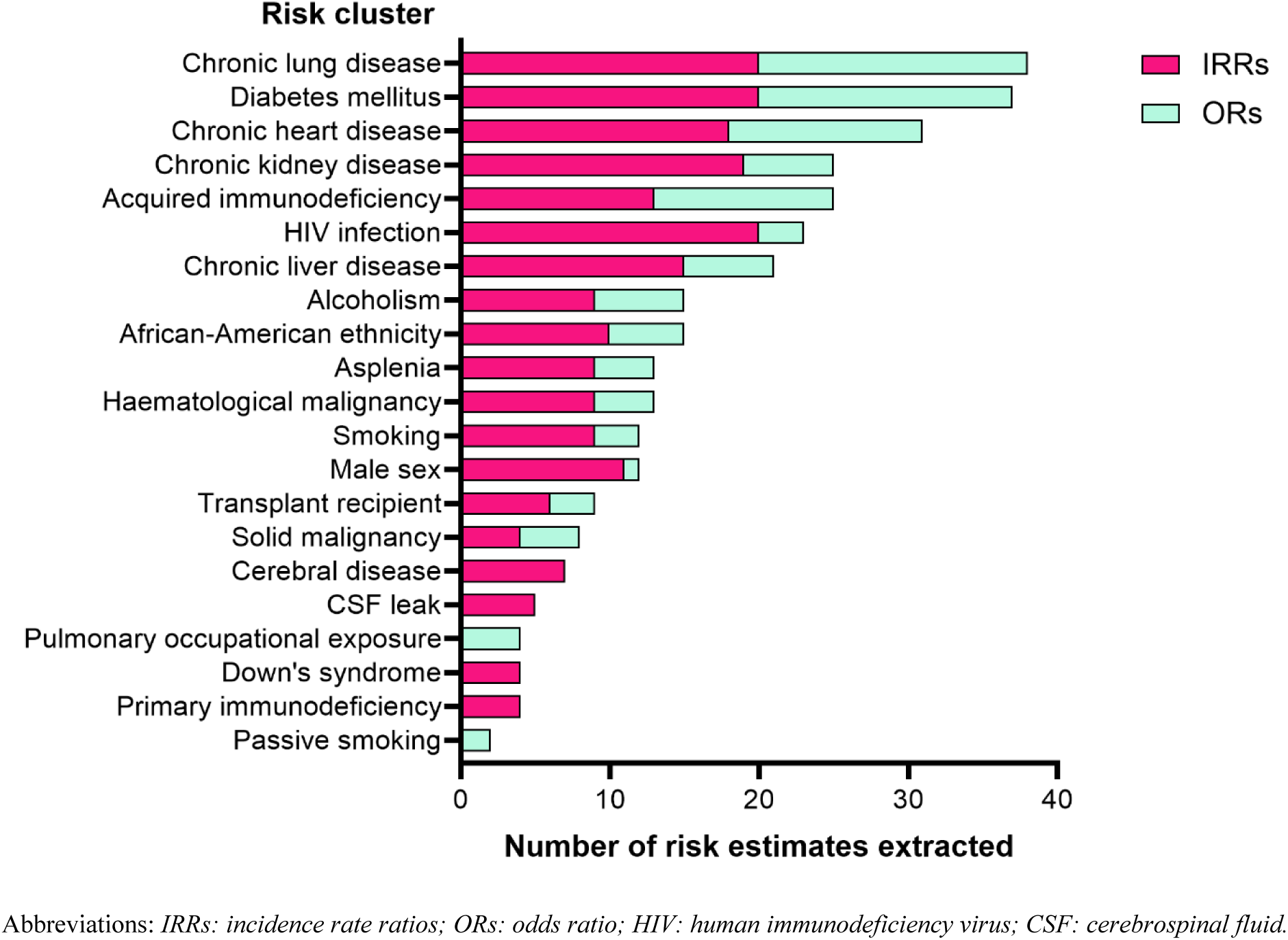
Number of risk estimates extracted per risk cluster. Data were extracted from 45 articles included in the meta-analyses.

The pooled risk estimates for reported IRRs for adult IPD are displayed in Figure 3. The pooled risk estimates for ORs are visualized in Supplementary material G. All individual forest plots that support the pooled risk estimates are provided in Supplementary material H and I, for IRRs and ORs respectively. For all 19 risk condition studied by IRRs, the level of the composite evidence was “very low” based on the GRADE assessment tool (Supplementary material J).

**Figure 3:**
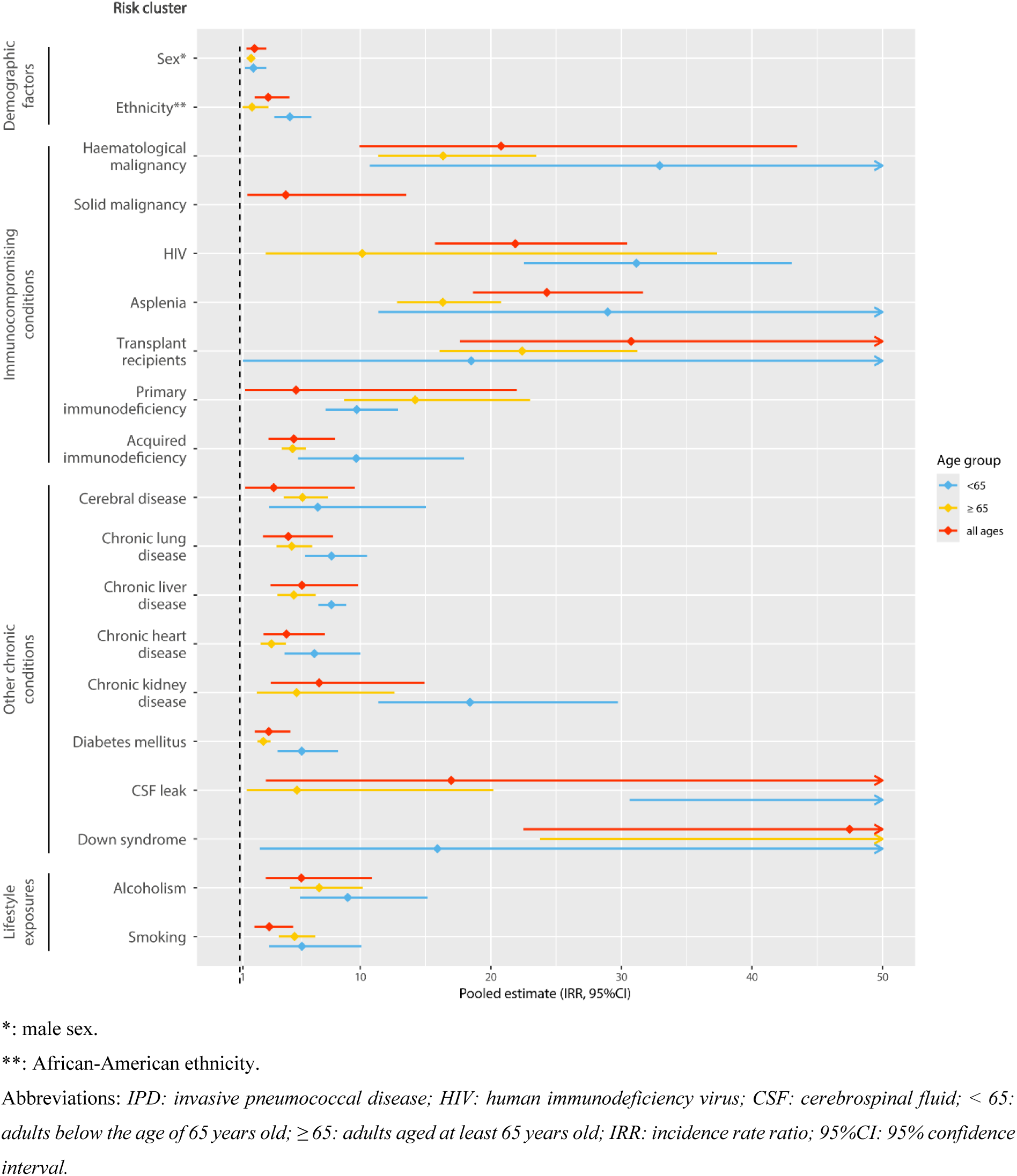
Pooled risk estimates of risk conditions for adult IPD. Summary of the studies that reported IRRs for the 19 risk clusters that were eligible for meta-analysis, stratified by age.

Risk clusters associated with the highest pooled risk estimates for adult IPD (IRRs > 10) were immunocompromising conditions such as haematological malignancy, HIV, transplant recipients and asplenia with an IRR of respectively 20·78 (95% CI 9·94-43·47), 21·87 (95% CI 15·72-30·43), 30·75 (95% CI 17·64-53·60) and 24·29 (95% CI 18·63-31·65). Other identified risk clusters with a high pooled risk estimate were compromised cerebrospinal fluid barrier (IRR 16·97, 95% CI 2·77-103.89) and Down’s syndrome (IRR 47·48, 95% CI 22·49-100·22).

Among other chronic conditions, kidney disease showed notably high IRRs for adult IPD, especially below the age of 65 with an IRR of 18·40 (95% CI 11·38-29·74). The smallest pooled IRR was observed for male sex (IRR 1·89, 95% CI 1·29-2·79). Age itself, after correction for the presence of other risk conditions, posed a relatively small direct risk for IPD (Supplementary material E). Rather, below the age of 65 the risk associated with a given condition was generally larger than in the older population, with significant increases observed for diabetes mellitus, African-American ethnicity, and chronic liver disease. The only exception to this pattern were younger patients with primary immunodeficiencies, in whom the IRR estimate for IPD was smaller (not significantly) than that of >65.

Risk conditions supported by single studies or otherwise unsuited for the current meta-analysis are listed in Table 2 and the extracted risk estimates are provided in Supplementary material K. The largest risk estimates were observed for Native American origin, Indigenous Australian origin, having lower income, living in a long-term care facility, and a history of a prior bacterial pneumonia. Looking at medication other than immunosuppressants, mainly immunostimulant medication (i.e. hematopoietic growth factors and interferons) were associated with an increased risk for IPD.

**Table 2:** Risk conditions not eligible for meta-analysis.

| Risk condition | Number of extracted risk estimates (See Supplementary material H) | Specified subtypes |
| --- | --- | --- |
| Ethnicity (other than African-American) | 10 | Aboriginal, Native American, Asian<sup>x</sup>, Hispanic<sup>x</sup> |
| Demography | 20 | Density<sup>x</sup>, % African American population, % population aged <5 or >65<sup>x</sup>, number of children living in household<sup>a</sup>, long-term care facility resident, town or village vs urban area<sup>x</sup> |
| Socio-economic factors | 30 | Homelessness, deprivation, unemployment, level of education, annual income |
| Weight | 5 | BMI <5<sup>th</sup> percentile<sup>x</sup>, BMI >95<sup>th</sup> percentile<sup>x</sup>, obesity |
| Medication usage (other than immunosuppressants) | 15 | Inhalation corticosteroids, opioids, immunostimulant drugs (i.e. hematopoietic growth factors and interferons), PPI or H2 blocker, anti-hypertensive medication<sup>x</sup>, anti-bacterial drugs |
| Coeliac disease<sup>b</sup> | 3 |  |
| Short gestation/low birthweight<sup>x</sup> | 1 |  |
| Occupational exposure (other than respiratory) | 13 | Day care worker<sup>x</sup>, farmer<sup>y</sup>, electrician<sup>x</sup>, amount of workers in close proximity, contact with coworkers/general public, regular contacts with infected patients<sup>x</sup>, sharing materials/surfaces with co-workers/general public, working outside versus inside, social distancing |
| Prior infection | 5 | Bacterial, influenza |
| Obstructive sleep apnoea | 3 |  |
| Metabolic disorders | 3 |  |

<sup>x</sup>not statistically significant.
<sup>a</sup>only children in household <6 years old in day care statistically significant.
<sup>b</sup>only statistically significant in patients aged 40-59,
<sup>y</sup>statistically significant lower risk of IPD
Abbreviations: *BMI*: body mass index; *PPI*: proton pump inhibitor; *H2*: histamine-2.

Four studies included in the meta-analyses assessed whether the number of combined risk conditions in an individual increases the overall risk of developing IPD. The types of counted risk conditions varied across studies, ranging from any condition, to specifically non-immunocompromising conditions. As shown in Figure 4, the IRRs for adult IPD were consistently higher in individuals with multimorbidity compared to those with a single risk condition.

**Figure 4:**
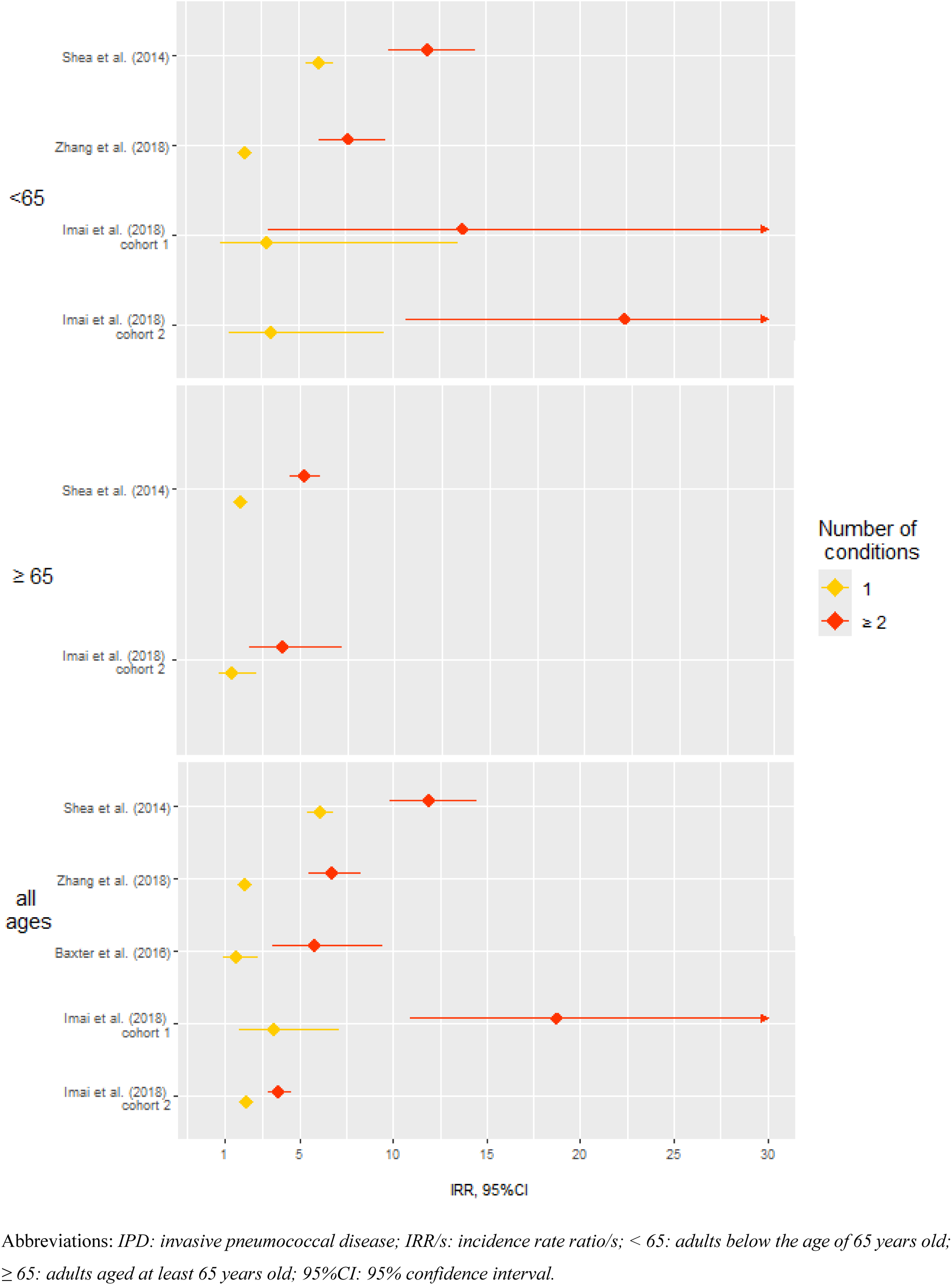
Risk estimates of multimorbidity for adult IPD. Forest plot of reported IRRs stratified by number of comorbidity risk conditions (color), over different age categories (vertical box).

## Discussion

This systematic review and meta-analyses provide the first comprehensive overview of all reported risk conditions for adult IPD and their relative importance. All 56 included studies had a high risk of bias, and the level composite evidence was very low for all risk conditions analysed. Meta-analyses were performed for 21 risk clusters, of which immunocompromising conditions, compromised cerebrospinal fluid barrier, Down’s syndrome, and chronic kidney disease were associated with the largest relative risk estimates for adult IPD. Below the age of 65 years old, virtually all studied risk conditions mediated a larger relative risk size, compared to in the older population. In addition, a limited number of studies reported on ethnic, demographic and socio-economic factors that may contribute to the risk of adult IPD. Adults with more than one risk condition displayed an accumulated risk for IPD.

To ensure capture of the variety of clinical manifestations that comprise IPD we applied a broad search strategy and performed iterative refinement until all eligible reports listed in relevant review articles were captured. The studies included in the systematic review were relatively old and lacked representation of certain populations like adults from low- and middle-income countries, patients with solid malignancies, and those receiving immunocompromising treatments. The studies included in the meta-analyses showed heterogeneity in study and control population and definition of the investigated risk conditions. Throughout the inclusion period the applied standard of care varied, which can affect the risk of IPD in terms of recognition and management of the underlying condition yet also via additional preventive measures like immunisation or pre-emptive use of antibiotics.^29,31^ This heterogeneity in combination with a high risk of bias were the main reasons for the overall very low quality of evidence. The validity of the pooled risk estimates for aggregated risk clusters may vary across sub-conditions. Detailed risk estimates for sub-conditions, singular studies, and ORs are provided in the Supplementary material. The main article features IRRs because the occurrence of IPD is relatively rare in all studied populations. In that circumstance, not accounting for follow-up years (yielding ORs) may have led to under- or overestimated attribution of captured IPD episodes to a certain risk condition, especially when the risk condition was rare as well.

The most pronounced risk conditions with large relative risk sizes were in line with the current indications for adult pneumococcal vaccination as recommended by the Advisory Committee on Immunization Practices.^79^ While in most adult IPD cohorts more than half of the cases is male,^80^ the relative risk size for IPD conferred by male sex was minor, as it was for smoking, African-American ethnicity, and diabetes mellitus. Secondary immunodeficiency, a risk cluster that included adults who use immunosuppressive medication, mediated a relative risk size that is comparable to those from chronic lung, liver, or heart disease or alcoholism. Conditions not eligible for meta-analysis but with relevant reported risk sizes for IPD were certain ethnicities, lower socio-economic status, and particularly homelessness. Although we excluded studies that reported risk factors for recurrent IPD, one study reported that preceding pneumonia conferred a significantly increased risk for IPD (IRR 5·5 and 2·9 for bacterial and influenza pneumonia respectively). The study that specifically focused on the risk profile for pneumococcal meningitis did not substantiate risk from chronic conditions other than past head or spinal injury.^81^

Studies that reported age category as risk condition for IPD had all applied adjustment for comorbidities. This indicates that while age may come with comorbidities, ageing in reasonable health only modestly increases one’s risk for IPD. The meta-analysis for the 21 risk clusters was performed differently, in that individuals with a risk condition were compared to members from the general population in the same age stratum. Therefore, the systematically larger relative risk sizes in the population below 65 years old, may fractionally be attributed to a relatively fit comparator population. While age-stratification reduced the between-study heterogeneity, IPD risks likely alter gradually with age which may be better appreciated by modelling instead of pre-stratification.

The relevance of multimorbidity leading to risk stacking was previously described for community acquired pneumonia.^82^ Our analysis confirms this phenomenon, showing substantial increases in IRRs for IPD in the presence of more than 1 risk condition, also among patients below the age of 65 years old.

Compared to current practices, populations that likely benefit from greater attention for prevention of IPD are the younger adults with risk conditions -especially chronic kidney disease, homeless individuals, and those with Down’s syndrome.

This systematic review and meta-analysis provides an overview that allows for comparison between populations at-risk for developing IPD. These findings may assist policy makers and clinicians in informing patients about their risk of IPD and in recommending or developing intervention strategies, such as vaccination, lifestyle changes, or optimal management of underlying conditions. The risk ratios that we report do not show the absolute risk size for IPD nor the size of the population at risk, which are both relevant to evaluate potential effectiveness of preventive interventions. Future research should focus on quantifying the cumulative risk associated with multiple concurrent risk conditions for developing IPD, as well as substantiating the relative efficacy of vaccination and other preventive measures in vulnerable populations.

## Author contribution

Study design: all authors. Literature search and data-extraction: KD and AC. Quality assessment: KD and AC. Statistical analysis: FG. First draft: KD. Review and editing: AC, JtO and FG.

## Data sharing

This study did not involve individual patient data, as the studied risk conditions were described by observational studies. All data extracted for this systematic review and used for the meta-analyses are provided in the Supplementary material.

## Data Availability

All data produced in the present work are contained in the manuscript.

## Acknowledgements

We kindly thank Alice Tillema from the Medical Library, Radboud University, The Netherlands for her help with the development of the search strategy.

## Supplementary material

### A. Search strategy and eligibility criteria for systematic review

**Databases:**

PubMed and Embase

Search performed on 03-04-2023 Updated search on 11-06-2024

#### Pubmed

(“Meningitis, Pneumococcal”[Mesh] OR invasive pneumococ*[tiab] OR “invasive pneumococcal disease”[Title/Abstract] OR “invasive pneumococcal pneumonia”[Title/Abstract] OR “pneumococcal meningitis”[Title/Abstract] OR “pneumococcal empyema”[Title/Abstract] OR “pneumococcal bacteraemi*”[Title/Abstract] OR “pneumococcal bacteremi*”[Title/Abstract] OR bacteremi* pneumococ*[tiab] OR bacteraemi* pneumococ*[tiab])

AND

(“risk factors”[MeSH Terms] OR “risk assessment”[MeSH Terms] OR “comorbidity”[MeSH Terms] OR “risk factor*”[Title/Abstract] OR “risk”[Title/Abstract] OR “at risk”[Title/Abstract] OR “at increased risk”[Title/Abstract] OR comorbid*[tiab] OR co-morbid*[tiab] OR “immunocompromised host”[MeSH Terms] OR “immunologic deficiency syndromes”[MeSH Terms] OR “immunocompromi*”[Title/Abstract] OR “immunodef*”[tiab])

Filters applied: English/Dutch language Updated search Pubmed:

#X AND (“2023/04/03”[CRDT] : “3000”[CRDT] OR “2024/04/03”[EDAT] : “3000”[EDAT] OR “2024/04/03”[MHDA] : “3000”[MHDA])

#### Embase

- (pneumococcal meningitis/ or invasive pneumococcal.ti,ab,kf. or “invasive pneumococcal disease”.ti,ab,kf. or “invasive pneumococcal pneumonia”.ti,ab,kf. or “pneumococcal meningitis”.ti,ab,kf. or “pneumococcal empyema”.ti,ab,kf. or “pneumococcal bacteraemia”.ti,ab,kf. or “pneumococcal bacteremia”.ti,ab,kf. or bacteremic pneumo*.ti,ab,kf. or bacteraemic pneomo*.ti,ab,kf.) and (Risk factor/ or exp risk assessment/ or comorbidity/ or “risk factor*”.ti,ab,kf. or “risk”.ti,ab,kf. or “at risk”.ti,ab,kf. or “at increased risk”.ti,ab,kf. or “comorbid*”.ti,ab,kf. or co-morbid*.ti,ab,kf. or immunocompromised patient/ or immunopathology/ or exp autoimmune disease/ or exp autoinflammatory disease/ or engraftment syndrome/ or exp graft rejection/ or exp graft versus host reaction/ or hyperinflammation/ or exp immune deficiency/ or immune dysregulation/ or immune injury/ or immune mediated injury/ or immune reconstitution inflammatory syndrome/ or exp paraproteinemia/ or “immunocompromi*”.ti,ab,kf. or “immunodef*”.ti,ab,kf.)
- limit 1 to (english language and “remove preprint records”)
- 2 and “Conference Abstract”.sa_pubt.
- 2 not 3

Updated search Embase:

limit 4 to dc=20230403-20240611

#### Inclusion criteria

- Reporting the risk of invasive pneumococcal disease in adults, in comparison to the general outpatient population

- Prevalence of risk factors is reported for the entire cohort

- Risk factors for invasive pneumococcal disease are described separately from risk factors for non-invasive pneumococcal pneumonia

#### Exclusion criteria

- Case reports

- Reviews not reporting primary data

- Studies regarding recurrent invasive pneumococcal disease

- Studies regarding host polymorphisms

- Studies in which >10% of the population concerns children (aged <18 years old)

- AMENDMENT: studies concerning children ≥14 years as ‘adults’ included

- Outcome measure not based on culture result or disease code

- No ratio of risk size reported

### B. List of excluded studies full text

#### Exclusion due to no relevant comparative measure

1. Zarabi N, Aldven M, Sjolander S, Wahl HF, Bencina G, Johnson KD, et al. Clinical and economic burden of pneumococcal disease among adults in Sweden: A population-based register study. PLoS ONE. 2023;18(7):e0287581.
2. Raman R, Brennan J, Ndi D, Sloan C, Markus TM, Schaffner W, et al. Marked Reduction of Socioeconomic and Racial Disparities in Invasive Pneumococcal Disease Associated with Conjugate Pneumococcal Vaccines. Journal of Infectious Diseases. 2021;223(7):1250-9.
3. Hyams C, Amin-Chowdhury Z, Fry NK, North P, Finn A, Judge A, et al. Streptococcus Pneumoniae septic arthritis in adults in Bristol and Bath, United Kingdom, 2006-2018: a 13-year retrospective observational cohort study. Emerging microbes & infections. 2021;10(1):1369-77.
4. Marrie TJ, Tyrrell GJ, Majumdar SR, Eurich DT. Effect of Age on the Manifestations and Outcomes of Invasive Pneumococcal Disease in Adults. The American journal of medicine. 2018;131(1):100.e1-.e7.
5. Marrie TJ, Tyrrell GJ, Majumdar SR, Eurich DT. Invasive Pneumococcal Disease: Still Lots to Learn and a Need for Standardized Data Collection Instruments. Canadian respiratory journal. 2017;2017:2397429.
6. Marrie TJ, Tyrrell GJ, Majumdar SR, Eurich DT. Concurrent Infection with Hepatitis C Virus and Streptococcus pneumoniae. Emerging infectious diseases. 2017;23(7):1118-23.
7. Fukusumi M, Chang B, Tanabe Y, Oshima K, Maruyama T, Watanabe H, et al. Invasive pneumococcal disease among adults in Japan, April 2013 to March 2015: disease characteristics and serotype distribution. BMC infectious diseases. 2017;17(1):2.
8. Chowers M, Regev-Yochay G, Mor O, Cohen-Poradosu R, Riesenberg K, Zimhony O, et al. Invasive pneumococcal disease (IPD) in HIV infected patients in Israel since the introduction of pneumococcal conjugated vaccines (PCV): Analysis of a nationwide surveillance study, 2009-2014. Human vaccines & immunotherapeutics. 2017;13(1):216-9.
9. Bareja C, Toms C, Lodo K, de Kluyver R. Invasive pneumococcal disease in Australia, 2009 and 2010. Australia2015 2015- 6-30. E265-79 p.
10. Yacoub AT, Monta R, Quaiser S, Acevedo I, Greene J. Pneumococcal Bacteremia in Patients with Cancer: A Retrospective Observational Study, 2003-2013. Infectious Diseases in Clinical Practice. 2015;23(5):263-6.
11. Wolter N, Tempia S, Cohen C, Madhi SA, Venter M, Moyes J, et al. High nasopharyngeal pneumococcal density, increased by viral coinfection, is associated with invasive pneumococcal pneumonia. The Journal of infectious diseases. 2014;210(10):1649-57.
12. Grau I, Ardanuy C, Calatayud L, Schulze MH, Liñares J, Pallares R. Smoking and alcohol abuse are the most preventable risk factors for invasive pneumonia and other pneumococcal infections. International journal of infectious diseases : IJID : official publication of the International Society for Infectious Diseases. 2014;25:59-64.
13. Morrill HJ, Caffrey AR, Noh E, LaPlante KL. Epidemiology of Pneumococcal Disease in a National Cohort of Older Adults. Infectious Diseases and Therapy. 2014;3(1):19-33.
14. Regev-Yochay G, Rahav G, Riesenberg K, Wiener-Well Y, Strahilevitz J, Stein M, et al. Initial effects of the National PCV7 Childhood Immunization Program on adult invasive pneumococcal disease in Israel. Plos one. 2014;9(2):e88406
15. Weinberger DM, Harboe ZB, Viboud C, Krause TG, Miller M, Mølbak K, et al. Pneumococcal disease seasonality: incidence, severity and the role of influenza activity. The European respiratory journal. 2014;43(3):833-41.
16. Muhammad RD, Oza-Frank R, Zell E, Link-Gelles R, Narayan KM, Schaffner W, et al. Epidemiology of invasive pneumococcal disease among high-risk adults since the introduction of pneumococcal conjugate vaccine for children. Clinical infectious diseases : an official publication of the Infectious Diseases Society of America. 2013;56(5):e59-67.
17. Gunaratnam PJ, Gilmour RE, Lowbridge C, McIntyre PB. Bug Breakfast in the Bulletin: invasive pneumococcal disease. New South Wales public health bulletin. 2013;24(3):142.
18. Fleming-Dutra KE, Taylor T, Link-Gelles R, Garg S, Jhung MA, Finelli L, et al. Effect of the 2009 influenza A(H1N1) pandemic on invasive pneumococcal pneumonia. The Journal of infectious diseases. 2013;207(7):1135-43.
19. Song JY, Choi JY, Lee JS, Bae IG, Kim YK, Sohn JW, et al. Clinical and economic burden of invasive pneumococcal disease in adults: a multicenter hospital-based study. BMC infectious diseases. 2013;13:202.
20. Regev-Yochay G, Rahav G, Strahilevitz J, Bishara J, Katzir M, Chowers M, et al. A nationwide surveillance of invasive pneumococcal disease in adults in Israel before an expected effect of PCV7. Vaccine. 2013;31(19):2387-94.
21. Weinberger DM, Harboe ZB, Viboud C, Krause TG, Miller M, Mølbak K, et al. Serotype-specific effect of influenza on adult invasive pneumococcal pneumonia. The Journal of infectious diseases. 2013;208(8):1274-80.
22. Rolo D, Fenoll A, Fontanals D, Larrosa N, Giménez M, Grau I, et al. Serotype 5 pneumococci causing invasive pneumococcal disease outbreaks in Barcelona, Spain (1997 to 2011). Journal of clinical microbiology. 2013;51(11):3585-90.
23. Rock C, Sadlier C, Fitzgerald J, Kelleher M, Dowling C, Kelly S, et al. Epidemiology of invasive pneumococcal disease and vaccine provision in a tertiary referral center. European journal of clinical microbiology & infectious diseases : official publication of the European Society of Clinical Microbiology. 2013;32(9):1135-41.
24. Christensen JS, Jensen TG, Kolmos HJ, Pedersen C, Lassen A. Bacteremia with Streptococcus pneumoniae: Sepsis and other risk factors for 30-day mortality-a hospital-based cohort study. European Journal of Clinical Microbiology and Infectious Diseases. 2012;31(10):2719-25.
25. Barry C, Krause VL, Cook HM, Menzies RI. Invasive pneumococcal disease in Australia 2007 and 2008. Communicable diseases intelligence quarterly report. 2012;36(2):E151-65.
26. Gharwan H, Gradon JD. Isolated pneumococcal bacteremia: Patients referred for infectious diseases consultation from 1999 to 2010. Infectious Diseases in Clinical Practice. 2011;19(1):34-7.
27. Gessner BD, Mueller JE, Yaro S. African meningitis belt pneumococcal disease epidemiology indicates a need for an effective serotype 1 containing vaccine, including for older children and adults. BMC infectious diseases. 2010;10:22.
28. . Rueda AM, Serpa JA, Matloobi M, Mushtaq M, Musher DM. The spectrum of invasive pneumococcal disease at an adult
29. tertiary care hospital in the early 21st century. Medicine. 2010;89(5):331-6.
30. Jansen AG, Rodenburg GD, de Greeff SC, Hak E, Veenhoven RH, Spanjaard L, et al. Invasive pneumococcal disease in the Netherlands: Syndromes, outcome and potential vaccine benefits. Vaccine. 2009;27(17):2394-401.
31. Plevneshi A, Svoboda T, Armstrong I, Tyrrell GJ, Miranda A, Green K, et al. Population-based surveillance for invasive pneumococcal disease in homeless adults in Toronto. PloS one. 2009;4(9):e7255.
32. Roche PW, Krause V, Cook H, Barralet J, Coleman D, Sweeny A, et al. Invasive pneumococcal disease in Australia, 2006. Communicable diseases intelligence quarterly report. 2008;32(1):18-30.
33. Bruce MG, Deeks SL, Zulz T, Bruden D, Navarro C, Lovgren M, et al. International circumpolar surveillance system for invasive pneumococcal disease, 1999-2005. Emerging Infectious Diseases. 2008;14(1):25-33.
34. Roche P, Krause V, Cook H, Bartlett M, Coleman D, Davis C, et al. Invasive pneumococcal disease in Australia, 2005. Communicable diseases intelligence quarterly report. 2007;31(1):86-100.
35. Chi R-C, Jackson LA, Neuzil KM. Characteristics and outcomes of older adults with community-acquired pneumococcal bacteremia. Journal of the American Geriatrics Society. 2006;54(1):115-20.
36. Greene CM, Kyaw MH, Ray SM, Schaffner W, Lynfield R, Barrett NL, et al. Preventability of invasive pneumococcal disease and assessment of current polysaccharide vaccine recommendations for adults: United States, 2001-2003. Clinical infectious diseases : an official publication of the Infectious Diseases Society of America. 2006;43(2):141-50.
37. Heffernan RT, Barrett NL, Gallagher KM, Hadler JL, Harrison LH, Reingold AL, et al. Declining incidence of invasive Streptococcus pneumoniae infections among persons with AIDS in an era of highly active antiretroviral therapy, 1995-2000. The Journal of infectious diseases. 2005;191(12):2038-45.
38. Reinert RR, Haupts S, van der Linden M, Heeg C, Cil MY, Al-Lahham A, et al. Invasive pneumococcal disease in adults in North-Rhine Westphalia, Germany, 2001-2003. Clinical Microbiology and Infection. 2005;11(12):985-91.
39. Shariatzadeh MR, Huang JQ, Tyrrell GJ, Johnson MM, Marrie TJ. Bacteremic pneumococcal pneumonia: A prospective study in Edmonton and neighboring municipalities. Medicine. 2005;84(3):147-61.
40. Jordano Q, Falcó V, Almirante B, Planes AM, del Valle O, Ribera E, et al. Invasive pneumococcal disease in patients infected with HIV: still a threat in the era of highly active antiretroviral therapy. Clinical infectious diseases : an official publication of the Infectious Diseases Society of America. 2004;38(11):1623-8.
41. Watt JP, O’Brien KL, Benin AL, Whitney CG, Robinson K, Parkinson AJ, et al. Invasive pneumococcal disease among Navajo adults, 1989-1998. Clinical infectious diseases : an official publication of the Infectious Diseases Society of America. 2004;38(4):496-501.
42. Kyaw MH, Christie P, Clarke SC, Mooney JD, Ahmed S, Jones IG, et al. Invasive Pneumococcal Disease in Scotland, 1999- 2001: Use of Record Linkage to Explore Associations between Patients and Disease in Relation to Future Vaccination Policy. Clinical Infectious Diseases. 2003;37(10):1283-91.
43. Laurichesse H, Romaszko JP, Nguyen LT, Souweine B, Poirier V, Guólon D, et al. Clinical characteristics and outcome of patients with invasive pneumococcal disease, Puy-de-Dôme, France, 1994-1998. European journal of clinical microbiology & infectious diseases : official publication of the European Society of Clinical Microbiology. 2001;20(5):299-308.
44. Gordon SB, Walsh AL, Chaponda M, Gordon MA, Soko D, Mbwvinji M, et al. Bacterial meningitis in Malawian adults: pneumococcal disease is common, severe, and seasonal. Clinical infectious diseases : an official publication of the Infectious Diseases Society of America. 2000;31(1):53-7.
45. McIntyre PB, Gilmour RE, Gilbert GL, Kakakios AM, Mellis CM. Epidemiology of invasive pneumococcal disease in urban New South Wales, 1997-1999. The Medical journal of Australia. 2000;173:S22-6.
46. Dworkin MS, Hanson DL. Epidemiologic relation between HIV and invasive pneumococcal disease in San Francisco County, California. Annals of internal medicine. 2000;132(12):1009.
47. Hogg GG, Strachan JE, Lester RA. Invasive pneumococcal disease in the population of Victoria. The Medical journal of Australia. 2000;173:S32-5.
48. Eber SW, Langendörfer CM, Ditzig M, Reinhardt D, Stöhr G, Soldan Wet al. Frequency of very late fatal sepsis after splenectomy for hereditary spherocytosis: impact of insufficient antibody response to pneumococcal infection. Germany; 1999 1999-11. Report No.: 0939-5555 (Print) Contract No.: 11.
49. Chen FM, Breiman RF, Farley M, Plikaytis B, Deaver K, Cetron MS. Geocoding and linking data from population-based surveillance and the US Census to evaluate the impact of median household income on the epidemiology of invasive Streptococcus pneumoniae infections. American journal of epidemiology. 1998;148(12):1212-8.
50. Campbell JF, Donohue MA, Mochizuki RB, Nevin-Woods CL, Spika JS. Pneumococcal bacteremia in Hawaii: initial findings of a pneumococcal disease prevention project. Hawaii medical journal. 1989;48(12):513-4, 7-8.
51. Simberkoff MS, El Sadr W, Schiffman G, Rahal JJ Jr. Streptococcus pneumoniae infections and bacteremia in patients with acquired immune deficiency syndrome, with report of a pneumococcal vaccine failure. United States; 1984 1984-12. Report No.: 0003-0805 (Print) Contract No.: 6.
52. Henneberger PK, Galaid EI, Marr JS. The descriptive epidemiology of pneumococcal meningitis in New York City. American journal of epidemiology. 1983;117(4):484-91.
53. Fraser DW, Darby CP, Koehler RE, Jacobs CF, Feldman RA. Risk factors in bacterial meningitis: Charleston County, South Carolina. The Journal of infectious diseases. 1973;127(3):271-7.

#### Exclusion due to no comparison to the general population

1. Nasreen S, Wang J, Marra F, Kwong JC, McGeer A, Sadarangani M, et al. Indirect impact of childhood 13-valent pneumococcal conjugate vaccine (PCV13) in Canadian older adults: A Canadian Immunization Research Network (CIRN) retrospective observational study. Thorax. 2024:e220377.
2. Calvo-Silveria S, Gonzalez-Diaz A, Grau I, Marimon JM, Cercenado E, Quesada MD, et al. Evolution of invasive pneumococcal disease by serotype 3 in adults: a Spanish three-decade retrospective study. The Lancet Regional Health - Europe. 2024;41:100913.
3. Liechti FD, Bijlsma MW, Brouwer MC, van Sorge NM, van de Beek D. Impact of the COVID-19 pandemic on incidence and serotype distribution of pneumococcal meningitis - A prospective, nationwide cohort study from the Netherlands. Journal of Infection. 2024;88(1):65-7.
4. Kang D-W, Kim C-R, Song JY, Park S-K. Cost-effectiveness of the 20-valent pneumococcal conjugate vaccine versus the 23-valent pneumococcal polysaccharide vaccine for older adults in South Korea. Vaccine. 2024;42(4):871-8.
5. Minassian D, Shan L, Dong C, Charania AN, Orihuela CJ, He C. Neighborhood-level disadvantages increase risk for invasive pneumococcal disease. American Journal of the Medical Sciences. 2024;367(5):304-9.
6. Anglemyer A, Ren X, Gilkison C, Kumbaroff Z, Morgan J, DuBray K, et al. The impact of pneumococcal serotype replacement on the effectiveness of a national immunization program: a population-based active surveillance cohort study in New Zealand. The Lancet Regional Health - Western Pacific. 2024;46:101082.
7. Amin-Chowdhury Z, Aiano F, Mensah A, Sheppard CL, Litt D, Fry NK, et al. Impact of the Coronavirus Disease 2019 (COVID-19) Pandemic on Invasive Pneumococcal Disease and Risk of Pneumococcal Coinfection With Severe Acute Respiratory Syndrome Coronavirus 2 (SARS-CoV-2): Prospective National Cohort Study, England. Clinical infectious diseases : an official publication of the Infectious Diseases Society of America. 2021;72(5):e65-e75.
8. Man MY, Shum HP, Yu JSY, Wu A, Yan WW. Burden of pneumococcal disease: 8-year retrospective analysis from a single centre in Hong Kong. Hong Kong medical journal = Xianggang yi xue za zhi. 2020;26(5):372-81.
9. Berry I, Tuite AR, Salomon A, Drews S, Harris AD, Hatchette T, et al. Association of Influenza Activity and Environmental Conditions With the Risk of Invasive Pneumococcal Disease. JAMA network open. 2020;3(7):e2010167.
10. Garcia Garrido HM, Mak AMR, Wit FWNM, Wong GWM, Knol MJ, Vollaard A, et al. Incidence and Risk Factors for Invasive Pneumococcal Disease and Community-acquired Pneumonia in Human Immunodeficiency Virus-Infected Individuals in a High-income Setting. Clinical infectious diseases : an official publication of the Infectious Diseases Society of America. 2020;71(1):41-50.
11. Willis R, Heslop O, Bodonaik N, Thame M, Smikle M. Morbidity, mortality and antimicrobial resistance of pneumococcal infections in the Jamaican paediatric and adult populations. Human antibodies. 2019;27(3):155-60.
12. de Celles MD, Arduin H, Levy-Bruhl D, Georges S, Souty C, Guillemot D, et al. Unraveling the seasonal epidemiology of pneumococcus. Proceedings of the National Academy of Sciences of the United States of America. 2019;116(5):1802-7.
13. Sadlier C, O’Connell S, Kelleher M, Bergin C. Incidence and risk factors for invasive pneumococcal disease in HIV-positive individuals in the era of highly active antiretroviral therapy. International journal of STD & AIDS. 2019;30(5):472-8.
14. Washio Y, Ito A, Kumagai S, Ishida T, Yamazaki A. A model for predicting bacteremia in patients with community-acquired pneumococcal pneumonia: a retrospective observational study. BMC pulmonary medicine. 2018;18(1):24.
15. de María Ugalde-Mejía L, Morales VA, Cárdenas G, Soto-Hernández JL. Adult Patients with Pneumococcal Meningitis at a Neurosurgical Neurologic Center: Different Predisposing Conditions? World neurosurgery. 2018;110:e642-e7.
16. Cowan J, Do TL, Desjardins S, Ramotar K, Corrales-Medina V, Cameron DW. Prevalence of Hypogammaglobulinemia in Adult Invasive Pneumococcal Disease. Clinical infectious diseases : an official publication of the Infectious Diseases Society of America. 2018;66(4):564-9.
17. Dalcin D, Sieswerda L, Dubois S, Ulanova M. Epidemiology of invasive pneumococcal disease in indigenous and non-indigenous adults in northwestern Ontario, Canada, 2006-2015. BMC infectious diseases. 2018;18(1):621.
18. Eton V, Schroeter A, Kelly L, Kirlew M, Tsang RSW, Ulanova M. Epidemiology of invasive pneumococcal and Haemophilus influenzae diseases in Northwestern Ontario, Canada, 2010-2015. International journal of infectious diseases : IJID : official publication of the International Society for Infectious Diseases. 2017;65:27-33.
19. Baldovin T, Lazzari R, Russo F, Bertoncello C, Buja A, Furlan P, et al. A surveillance system of Invasive Pneumococcal Disease in North-Eastern Italy. Annali di igiene : medicina preventiva e di comunita. 2016;28(1):15-24.
20. Medeiros MI, Negrini BV, Silva JM, Almeida SC, Leopoldo ML, Leopoldo Silva Guerra ML, et al. Clinical and microbiological implications of invasive pneumococcal disease in hospitalized patients (1998-2013). The Brazilian journal of infectious diseases : an official publication of the Brazilian Society of Infectious Diseases. 2016;20(3):242-9.
21. Cabaj JL, Nettel-Aguirre A, MacDonald J, Vanderkooi OG, Kellner JD. Influence of Childhood Pneumococcal Conjugate Vaccines on Invasive Pneumococcal Disease in Adults With Underlying Comorbidities in Calgary, Alberta (2000-2013). Clinical infectious diseases : an official publication of the Infectious Diseases Society of America. 2016;62(12):1521-6.
22. Toms C, de Kluyver R. Invasive pneumococcal disease in Australia, 2011 and 2012. Communicable diseases intelligence quarterly report. 2016;40(2); E267-84.
23. Askim Å, Mehl A, Paulsen J, DeWan AT, Vestrheim DF, Åsvold BO, et al. Epidemiology and outcome of sepsis in adult patients with Streptococcus pneumoniae infection in a Norwegian county 1993-2011: an observational study. BMC infectious diseases. 2016;16:223.
24. Burgos J, Larrosa MN, Martinez A, Belmonte J, González-López J, Rello J, et al. Impact of influenza season and environmental factors on the clinical presentation and outcome of invasive pneumococcal disease. European journal of clinical microbiology & infectious diseases : official publication of the European Society of Clinical Microbiology. 2015;34(1):177-86.
25. Pedro-Botet ML, Burgos J, Luján M, Gimenez M, Rello J, Planes A, et al. Impact of the 2009 influenza A H1N1 pandemic on invasive pneumococcal disease in adults. Scandinavian journal of infectious diseases. 2014;46(3):185-92.
26. Belkhir L, Rodriguez-Villalobos H, Vandercam B, Marot JC, Cornu O, Lambert M, et al. Pneumococcal septic arthritis in adults: clinical analysis and review. Acta clinica Belgica. 2014;69(1):40-6.
27. Verhaegen J, Flamaing J, De Backer W, Delaere B, Van Herck K, Surmont F, et al. Epidemiology and outcome of invasive pneumococcal disease among adults in Belgium, 2009-2011. Euro surveillance : bulletin Europeen sur les maladies transmissibles = European communicable disease bulletin. 2014;19(31):14-22.
28. Munier AL, de Lastours V, Porcher R, Donay JL, Pons JL, Molina JM. Risk factors for invasive pneumococcal disease in HIV-infected adults in France in the highly active antiretroviral therapy era. International journal of STD & AIDS.
29. Torda A, Chong Q, Lee A, Chen S, Dodds A, Greenwood M, et al. Invasive pneumococcal disease following adult allogeneic hematopoietic stem cell transplantation. Transplant infectious disease : an official journal of the Transplantation Society. 2014;16(5):751-9.
30. Seminog OO, Goldacre MJ. Risk of pneumonia and pneumococcal disease in people hospitalized with diabetes mellitus: English record-linkage studies. England2013 2013-12. 1412-9 p.
31. Kang CI, Song JH, Kim SH, Chung DR, Peck KR, Thamlikitkul V, et al. Risk factors and pathogenic significance of bacteremic pneumonia in adult patients with community-acquired pneumococcal pneumonia. The Journal of infection. 2013;66(1):34-40.
32. Yin Z, Rice BD, Waight P, Miller E, George R, Brown AE, et al. Invasive pneumococcal disease among HIV-positive individuals, 2000-2009. AIDS (London, England). 2012;26(1):87-94.
33. Burgos J, Peñaranda M, Payeras A, Villoslada A, Curran A, Garau M, et al. Invasive pneumococcal disease in HIV-infected adults: clinical changes after the introduction of the pneumococcal conjugate vaccine in children. Journal of acquired immune deficiency syndromes (1999). 2012;59(1):31-8.
34. Grau I, Ardanuy C, Calatayud L, Rolo D, Domenech A, Liñares J, et al. Invasive pneumococcal disease in healthy adults: increase of empyema associated with the clonal-type Sweden(1)-ST306. PloS one. 2012;7(8):e42595.
35. van Deursen AM, van Mens SP, Sanders EA, Vlaminckx BJ, de Melker HE, Schouls LM, et al. Invasive pneumococcal disease and 7-valent pneumococcal conjugate vaccine, the Netherlands. Emerging infectious diseases. 2012;18(11):1729-37.
36. Kuster SP, Tuite AR, Kwong JC, McGeer A, Fisman DN. Evaluation of coseasonality of influenza and invasive pneumococcal disease: results from prospective surveillance. PLoS medicine. 2011;8(6):e1001042.
37. Siemieniuk RA, Gregson DB, Gill MJ. The persisting burden of invasive pneumococcal disease in HIV patients: an observational cohort study. BMC infectious diseases. 2011;11:314.
38. Nunes MC, von Gottberg A, de Gouveia L, Cohen C, Kuwanda L, Karstaedt AS, et al. Persistent high burden of invasive pneumococcal disease in South African HIV-infected adults in the era of an antiretroviral treatment program. PloS one. 2011;6(11):e27929.
39. Thigpen MC, Whitney CG, Messonnier NE, Zell ER, Lynfield R, Hadler JL, et al. Bacterial meningitis in the United States, 1998-2007. New England Journal of Medicine. 2011;364(21):2016-25.
40. Burgos J, Lujan M, Falcó V, Sánchez A, Puig M, Borrego A, et al. The spectrum of pneumococcal empyema in adults in the early 21st century. Clinical infectious diseases : an official publication of the Infectious Diseases Society of America. 2011;53(3):254-61.
41. Burckhardt I, Burckhardt F, van der Linden M, Heeg C, Reinert RR. Risk factor analysis for pneumococcal meningitis in adults with invasive pneumococcal infection. Epidemiology and infection. 2010;138(9):1353-8.
42. Cohen AL, Harrison LH, Farley MM, Reingold AL, Hadler J, Schaffner W, et al. Prevention of invasive pneumococcal disease among HIV-infected adults in the era of childhood pneumococcal immunization. AIDS (London, England). 2010;24(14):2253-62.
43. Namayanja-Kaye GA, Namale A, Joloba ML, Salata RA. Outcome of patients with pneumococcal bacteremia at Mulago Hospital, Kampala. Infectious Diseases in Clinical Practice. 2009;17(4):248-52.
44. White AN, Ng V, Spain CV, Johnson CC, Kinlin LM, Fisman DN. Let the sun shine in: effects of ultraviolet radiation on invasive pneumococcal disease risk in Philadelphia, Pennsylvania. BMC infectious diseases. 2009;9:196.
45. Peñaranda M, Falco V, Payeras A, Jordano Q, Curran A, Pareja A, et al. Effectiveness of polysaccharide pneumococcal vaccine in HIV-infected patients: a case-control study. Clinical infectious diseases : an official publication of the Infectious Diseases Society of America. 2007;45(7):e82-7.
46. Benca J, Lesnakova A, Holeckova K, Ondrusova A, Wiczmandyova O, Sladecko V, et al. Pneumococcal meningitis in community is frequent after craniocerebral trauma and in alcohol abusers. Sweden2007 2007-11. 16-7 p.
47. Parsons HK, Metcalf SC, Tomlin K, Read RC, Dockrell DH. Invasive pneumococcal disease and the potential for prevention by vaccination in the United Kingdom. The Journal of infection. 2007;54(5):435-8.
48. Barry PM, Zetola N, Keruly JC, Moore RD, Gebo KA, Lucas GM. Invasive pneumococcal disease in a cohort of HIV-infected adults: incidence and risk factors, 1990-2003. AIDS (London, England). 2006;20(3):437-44.
49. Watson M, Gilmour R, Menzies R, Ferson M, McIntyre P. The association of respiratory viruses, temperature, and other climatic parameters with the incidence of invasive pneumococcal disease in Sydney, Australia. Clinical infectious diseases : an official publication of the Infectious Diseases Society of America. 2006;42(2):211-5.
50. Weisfelt M, van de Beek D, Spanjaard L, Reitsma JB, de Gans J. Clinical features, complications, and outcome in adults with pneumococcal meningitis: a prospective case series. The Lancet Neurology. 2006;5(2):123-9.
51. Grabowska K, Högberg L, Penttinen P, Svensson A, Ekdahl K. Occurrence of invasive pneumococcal disease and number of excess cases due to influenza. BMC infectious diseases. 2006;6:58.
52. Kumashi P, Girgawy E, Tarrand JJ, Rolston KV, Raad II, Safdar A. Streptococcus pneumoniae bacteremia in patients with cancer: disease characteristics and outcomes in the era of escalating drug resistance (1998-2002). Medicine. 2005;84(5):303- 12.
53. Grau I, Pallares R, Tubau F, Schulze MH, Llopis F, Podzamczer D, et al. Epidemiologic changes in bacteremic pneumococcal disease in patients with human immunodeficiency virus in the era of highly active antiretroviral therapy. United States2005 2005-7-11. 1533-40 p.
54. Engelhard D, Cordonnier C, Shaw PJ, Parkalli T, Guenther C, Martino R, et al. Early and late invasive pneumococcal infection following stem cell transplantation: A European Bone Marrow Transplantation survey. British Journal of Haematology. 2002;117(2):444-50.
55. Burman LA, Norrby R, Trollfors B. Invasive pneumococcal infections: incidence, predisposing factors, and prognosis. Reviews of infectious diseases. 1985;7(2):133-42.
56. Chou MY, Brown AE, Blevins A, Armstrong D. Severe pneumococcal infection in patients with neoplastic disease. Cancer.

#### Exclusion due to not containing primary data

1. Lee MS. Invasive Pneumococcal Diseases in Korean Adults After the Introduction of Pneumococcal Vaccine into the National Immunization Program. Infection and Chemotherapy. 2023;55(4):411-21.
2. Campling J, Vyse A, Liu H-H, Wright H, Slack M, Reinert R-R, et al. A review of evidence for pneumococcal vaccination in adults at increased risk of pneumococcal disease: risk group definitions and optimization of vaccination coverage in the United Kingdom. Expert Review of Vaccines. 2023;22(1):785-800.
3. Ishiwada N. Current situation and need for prevention of invasive pneumococcal disease and pneumococcal pneumonia in 6- to 64-year-olds in Japan. Journal of infection and chemotherapy : official journal of the Japan Society of Chemotherapy. 2021;27(1):7-18.
4. Harris JG. Improving Pneumococcal Vaccination Rates in Rheumatology Clinics. The Journal of rheumatology. 2021.
5. Im H, Ser J, Sim U, Cho H. Promising Expectations for Pneumococcal Vaccination during COVID-19. Vaccines. 2021;9(12):1507.
6. Tyrrell G, Lee C, Eurich D. Is there a need for pneumococcal vaccination programs for the homeless to prevent invasive pneumococcal disease? Expert review of vaccines. 2021;20(9):1113-21.
7. Navarro-Torne A, Montuori EA, Kossyvaki V, Mendez C. Burden of pneumococcal disease among adults in Southern Europe (Spain, Portugal, Italy, and Greece): a systematic review and meta-analysis. Human Vaccines and Immunotherapeutics. 2021;17(10):3670-86.
8. Holzer L, Hoffman T, Van Kessel DA, Rijkers GT. Pneumococcal vaccination in lung transplant patients. Expert review of vaccines. 2020;19(3):227-34.
9. Dernoncourt A, El Samad Y, Schmidt J, Emond JP, Gouraud C, Brocard A, et al. Case studies and literature review of pneumococcal septic arthritis in adults. Emerging Infectious Diseases. 2019;25(10):1824-33.
10. van Aalst M, Lötsch F, Spijker R, van der Meer JTM, Langendam MW, Goorhuis A, et al. Incidence of invasive pneumococcal disease in immunocompromised patients: A systematic review and meta-analysis. Travel medicine and infectious disease. 2018;24:89-100.
11. Ceyhan M, Dagan R, Sayiner A, Chernyshova L, Dinleyici EC, Hryniewicz W, et al. Surveillance of pneumococcal diseases in Central and Eastern Europe. Human Vaccines and Immunotherapeutics. 2016;12(8):2124-34.
12. Chalmers JD, Campling J, Dicker A, Woodhead M, Madhava H. A systematic review of the burden of vaccine preventable pneumococcal disease in UK adults. BMC Pulmonary Medicine. 2016;16(1):77.
13. Curcio D, Cané A, Isturiz R. Redefining risk categories for pneumococcal disease in adults: critical analysis of the evidence. International journal of infectious diseases : IJID : official publication of the International Society for Infectious Diseases. 2015;37:30-5.
14. Torres A, Blasi F, Dartois N, Akova M. Which individuals are at increased risk of pneumococcal disease and why? Impact of COPD, asthma, smoking, diabetes, and/or chronic heart disease on community-acquired pneumonia and invasive pneumococcal disease. Thorax. 2015;70(10):984-9.
15. Drijkoningen JJ, Rohde GG. Pneumococcal infection in adults: burden of disease. Clinical microbiology and infection : the official publication of the European Society of Clinical Microbiology and Infectious Diseases. 2014;20:45-51.
16. Cruickshank HC, Jefferies JM, Clarke SC. Lifestyle risk factors for invasive pneumococcal disease: a systematic review. BMJ open. 2014;4(6):e005224.
17. Ash SY, Sheffield JVL. Pneumococcus. Medical Clinics of North America. 2013;97(4):647-66.
18. Hung IF, Tantawichien T, Tsai YH, Patil S, Zotomayor R. Regional epidemiology of invasive pneumococcal disease in Asian adults: epidemiology, disease burden, serotype distribution, and antimicrobial resistance patterns and prevention. International journal of infectious diseases : IJID : official publication of the International Society for Infectious Diseases. 2013;17(6):e364-73.
19. Boikos C, Quach C. Risk of invasive pneumococcal disease in children and adults with asthma: a systematic review. Vaccine. 2013;31(42):4820-6.
20. Amoateng-Adjepong Y. Is sickle cell trait a risk factor for invasive pneumococcal disease? United States2010 2010-5. 347-8 p.
21. Prommalikit O, Pengsaa K, Thisyakorn U. Pneumococcal infections in high-risk and immunocompromised hosts. Journal of the Medical Association of Thailand = Chotmaihet thangphaet. 2010;93:S61-70.
22. Wei BP, Shepherd RK, Robins-Browne RM, Clark GM, O’Leary SJ. Pneumococcal meningitis post-cochlear implantation: potential routes of infection and pathophysiology. Otolaryngology--head and neck surgery : official journal of American Academy of Otolaryngology-Head and Neck Surgery. 2010;143(5):S15-23.
23. Wei BP, Shepherd RK, Robins-Browne RM, Clark GM, O’Leary SJ. Pneumococcal meningitis post-cochlear implantation: preventative measures. Otolaryngology--head and neck surgery : official journal of American Academy of Otolaryngology-Head and Neck Surgery. 2010;143(5):S9-14.
24. Juhn YJ. Asthma and invasive pneumococccal diseases: the implications for clinical practice, public health and research. Paediatrics and Child Health. 2009;19:S127-S31.
25. Lynch JP 3rd, Zhanel GG. Streptococcus pneumoniae: epidemiology, risk factors, and strategies for prevention. Seminars in respiratory and critical care medicine. 2009;30(2):189-209.
26. Lexau CA. The changing epidemiology of pneumococcal pulmonary disease in the era of the heptavalent vaccine. Current Infectious Disease Reports. 2008;10(3):229-35.
27. Fletcher MA, Laufer DS, McIntosh ED, Cimino C, Malinoski FJ. Controlling invasive pneumococcal disease: is vaccination of at-risk groups sufficient? International journal of clinical practice. 2006;60(4):450-6.
28. Cotton D, Kuschner WG, Kuschner RA, Talbot TR, Hartert TV, Griffin MR. Asthma and invasive pneumococcal disease [5] (multiple letters). New England Journal of Medicine. 2005;353(7):738-9.
29. Ortqvist A, Hedlund J, Kalin M. Streptococcus pneumoniae: epidemiology, risk factors, and clinical features. Seminars in
30. Taylor SN, Sanders CV. Unusual manifestations of invasive pneumococcal infection. The American journal of medicine. 1999;107(1):12S-27S.
31. Van Beneden C. Epidemiology of pneumococcal disease. American Journal of Managed Care. 1999;5:S991-S9.
32. Janoff EN, Rubins JB. Invasive pneumococcal disease in the immunocompromised host. Microbial drug resistance (Larchmont, NY). 1997;3(3):215-32.

#### Exclusion due to paediatric population

1. Navalkele P, Özgönenel B, McGrath E, Lephart P, Sarnaik S. Invasive Pneumococcal Disease in Patients With Sickle Cell Disease. Journal of pediatric hematology/oncology. 2017;39(5):341-4.
2. Wortham JM, Zell ER, Pondo T, Harrison LH, Schaffner W, Lynfield R, et al. Racial disparities in invasive streptococcus pneumoniae infections, 1998-2009. Clinical Infectious Diseases. 2014;58(9):1250-7.
3. Edwards LJ, Markey PG, Cook HM, Trauer JM, Krause VL. The relationship between influenza and invasive pneumococcal disease in the Northern Territory, 2005-2009. Medical Journal of Australia. 2011;194(4):207.
4. Poehling KA, Light LS, Rhodes M, Snively BM, Halasa NB, Mitchel E, et al. Sickle cell trait, hemoglobin C trait, and invasive pneumococcal disease. Epidemiology (Cambridge, Mass). 2010;21(3):340-6.
5. Rodenburg GD, de Greeff SC, Jansen AGCS, de Melker HE, Schouls LM, Hak E, et al. Effects of pneumococcal conjugate vaccine 2 years after its introduction, the Netherlands. Emerging Infectious Diseases. 2010;16(5):816-23.
6. Steenhoff AP, Wood SM, Rutstein RM, Wahl A, McGowan KL, Shah SS. Invasive pneumococcal disease among human immunodeficiency virus-infected children, 1989-2006. The Pediatric infectious disease journal. 2008;27(10):886-91.
7. Kyaw MH, Clarke S, Jones IG, Campbell H. Incidence of invasive pneumococcal disease in Scotland, 1988-99. Epidemiology and infection. 2002;128(2):139-47.
8. Robinson KA, Baughman W, Rothrock G, Barrett NL, Pass M, Lexau C, et al. Epidemiology of invasive Streptococcus pneumoniae infections in the United States, 1995-1998: Opportunities for prevention in the conjugate vaccine era. JAMA. 2001;285(13):1729-35.
9. Krause VL, Reid SJ, Merianos A. Invasive pneumococcal disease in the Northern Territory of Australia, 1994-1998. The Medical journal of Australia. 2000;173:S27-31.
10. The Vaccine Impact Surveillance Network--Invasive Pneumococcal Study Group. Are current recommendations for pneumococcal vaccination appropriate for Western Australia? The Medical journal of Australia. 2000;173:S36-40.
11. Pastor P, Medley F, Murphy TV. Invasive pneumococcal disease in Dallas County, Texas: results from population-based surveillance in 1995. Clinical infectious diseases : an official publication of the Infectious Diseases Society of America. 1998;26(3):590-5.
12. Trotman J, Hughes B, Mollison L. Invasive pneumococcal disease in central Australia. Clinical infectious diseases : an official publication of the Infectious Diseases Society of America. 1995;20(6):1553-6.

#### Exclusion due to study of host genetics

1. Kloek AT, Brouwer MC, van de Beek D. Host genetic variability and pneumococcal disease: a systematic review and meta-analysis. BMC medical genomics. 2019;12(1):130.
2. Sangil A, Arranz MJ, Güerri-Fernández R, Pérez M, Monzón H, Payeras A, et al. Genetic susceptibility to invasive pneumococcal disease. Infection, genetics and evolution : journal of molecular epidemiology and evolutionary genetics in infectious diseases. 2018;59:126-31.
3. Lingappa JR, Dumitrescu L, Zimmer SM, Lynfield R, McNicholl JM, Messonnier NE, et al. Identifying host genetic risk factors in the context of public health surveillance for invasive pneumococcal disease. PloS one. 2011;6(8):e23413.
4. Chapman SJ, Khor CC, Vannberg FO, Rautanen A, Walley A, Segal S, et al. Common NFKBIL2 polymorphisms and susceptibility to pneumococcal disease: a genetic association study2010 2010. R227 p.
5. Horcajada JP, Lozano F, Muñoz A, Suarez B, Fariñas-Alvarez C, Almela M, et al. Polymorphic receptors of the innate immune system (MBL/MASP-2 and TLR2/4) and susceptibility to pneumococcal bacteremia in HIV-infected patients: a case-control study. Current HIV research. 2009;7(2):218-23.
6. Payton A, Payne D, Mankhambo LA, Banda DL, Hart CA, Ollier WE, et al. Nitric oxide synthase 2A (NOS2A) polymorphisms are not associated with invasive pneumococcal disease2009 2009-3-23. 28 p.
7. Hjuler T, Poulsen G, Wohlfahrt J, Kaltoft M, Biggar RJ, Melbye M. Genetic susceptibility to severe infection in families with invasive pneumococcal disease. American journal of epidemiology. 2008;167(7):814-9.
8. Chapman SJ, Khor CC, Vannberg FO, Frodsham A, Walley A, Maskell NA, et al. IkappaB genetic polymorphisms and invasive pneumococcal disease. American journal of respiratory and critical care medicine. 2007;176(2):181-7.
9. Moens L, Van Hoeyveld E, Verhaegen J, De Boeck K, Peetermans WE, Bossuyt X. Fcgamma-receptor IIA genotype and invasive pneumococcal infection. Clinical immunology (Orlando, Fla). 2006;118(1):20-3.
10. Roy S, Knox K, Segal S, Griffiths D, Moore CE, Welsh KI, et al. MBL genotype and risk of invasive pneumococcal disease: a case-control study. Lancet (London, England). 2002;359(9317):1569-73.
11. Kronborg G, Weis N, Madsen HO, Pedersen SS, Wejse C, Nielsen H, et al. Variant mannose-binding lectin alleles are not associated with susceptibility to or outcome of invasive pneumococcal infection in randomly included patients. The Journal of infectious diseases. 2002;185(10):1517-20.

#### Exclusion due to combined risk of community-acquired pneumonia and invasive pneumococcal disease

1. Ochoa-Gondar O, Torras-Vives V, de Diego-Cabanes C, Satue-Gracia EM, Vila-Rovira A, Forcadell-Perisa MJ, et al. Incidence and risk factors of pneumococcal pneumonia in adults: a population-based study. BMC Pulmonary Medicine. 2023;23(1):200.
2. Kang JM, Kim EH, Ihn K, Jung I, Han M, Ahn JG. Risk of invasive pneumococcal disease in patients with
3. Seminog OO, Goldacre MJ. Risk of pneumonia and pneumococcal disease in people with severe mental illness: English record linkage studies. Thorax. 2013;68(2):171-6.
4. Wotton CJ, Goldacre MJ. Risk of invasive pneumococcal disease in people admitted to hospital with selected immune-mediated diseases: record linkage cohort analyses. Journal of epidemiology and community health. 2012;66(12):1177-81.
5. Juhn YJ, Kita H, Yawn BP, Boyce TG, Yoo KH, McGree ME, et al. Increased risk of serious pneumococcal disease in patients with asthma. The Journal of allergy and clinical immunology. 2008;122(4):719-23.
6. Thomas HJ, Wotton CJ, Yeates D, Ahmad T, Jewell DP, Goldacre MJ. Pneumococcal infection in patients with coeliac disease. European journal of gastroenterology & hepatology. 2008;20(7):624-8.
7. Kulkarni S, Powles R, Treleaven J, Riley U, Singhal S, Horton C, et al. Chronic graft versus host disease is associated with long-term risk for pneumococcal infections in recipients of bone marrow transplants. Blood. 2000;95(12):3683-6.
8. Gilks CF, Ojoo SA, Ojoo JC, Brindle RJ, Paul J, Batchelor BI, et al. Invasive pneumococcal disease in a cohort of predominantly HIV-1 infected female sex-workers in Nairobi, Kenya. Lancet (London, England). 1996;347(9003):718-23.
9. Hoge CW, Reichler MR, Dominguez EA, Bremer JC, Mastro TD, Hendricks KA, et al. An epidemic of pneumococcal disease in an overcrowded, inadequately ventilated jail. The New England journal of medicine. 1994;331(10):643-8.

#### Exclusion due to study of recurrent IPD

1. Malo JA, Ware RS, Lambert SB. Estimating the risk of recurrent invasive pneumococcal disease in Australia, 1991-2016. Vaccine. 2021;39(40):5748-56.

#### Exclusion due to no English language

1. Petrousova L, Roznovsky L, Maresova V. Pneumococcal meningitides at the clinic of infectious diseases in the university hospital in ostrava in 2004-2012. Vakcinologie. 2013;7(2):58-61.

#### Exclusion due to duplicate population

1. Torén K, Blanc PD, Qvarfordt I, Aspevall O, Schiöler L. Inhaled Corticosteroids Use and Risk of Invasive Pneumococcal Disease in a Population-based Study. Annals of the American Thoracic Society. 2020;17(12):1570-5.
2. Chu V, Carpenter DM, Winter K, Harriman K, Glaser C. Increased Risk of Late-onset Streptococcus pneumoniae Meningitis in Adults with Prior Head or Spine Surgeries. Clinical Infectious Diseases. 2019;68(12):2120-2.
3. Pelton SI, Bornheimer R, Doroff R, Shea KM, Sato R, Weycker D. Decline in Pneumococcal Disease Attenuated in Older Adults and Those With Comorbidities Following Universal Childhood PCV13 Immunization. Clinical infectious diseases : an official publication of the Infectious Diseases Society of America. 2019;68(11):1831-8.
4. Mosites E, Zulz T, Bruden D, Nolen L, Frick A, Castrodale L, et al. Risk for invasive streptococcal infections among adults experiencing homelessness, anchorage, Alaska, USA, 2002-2015. Emerging Infectious Diseases. 2019;25(10):1903-10.
5. Marcus JL, Baxter R, Leyden WA, Muthulingam D, Yee A, Horberg MA, et al. Invasive Pneumococcal Disease Among HIV-Infected and HIV-Uninfected Adults in a Large Integrated Healthcare System. AIDS patient care and STDs. 2016;30(10):463-70.
6. Ludvigsson JF, Olén O, Bell M, Ekbom A, Montgomery SM. Coeliac disease and risk of sepsis. Gut. 2008;57(8):1074-80.

### C. Global distribution of included studies

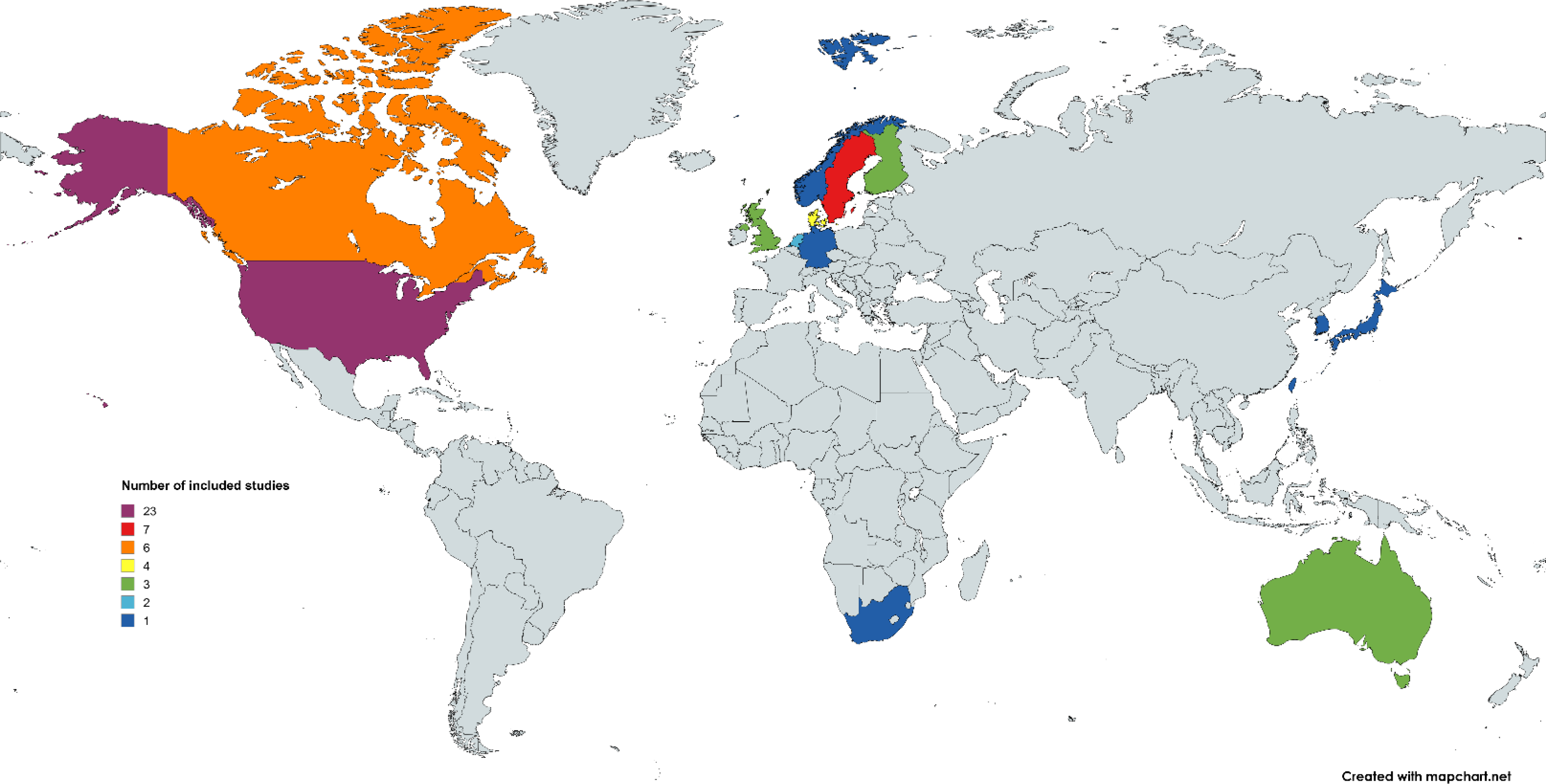

Global country map demonstrates the location and number (colour) of studies included in the systematic review. One study included data from both Canada as well as the United States.

### D. Risk of bias assessment

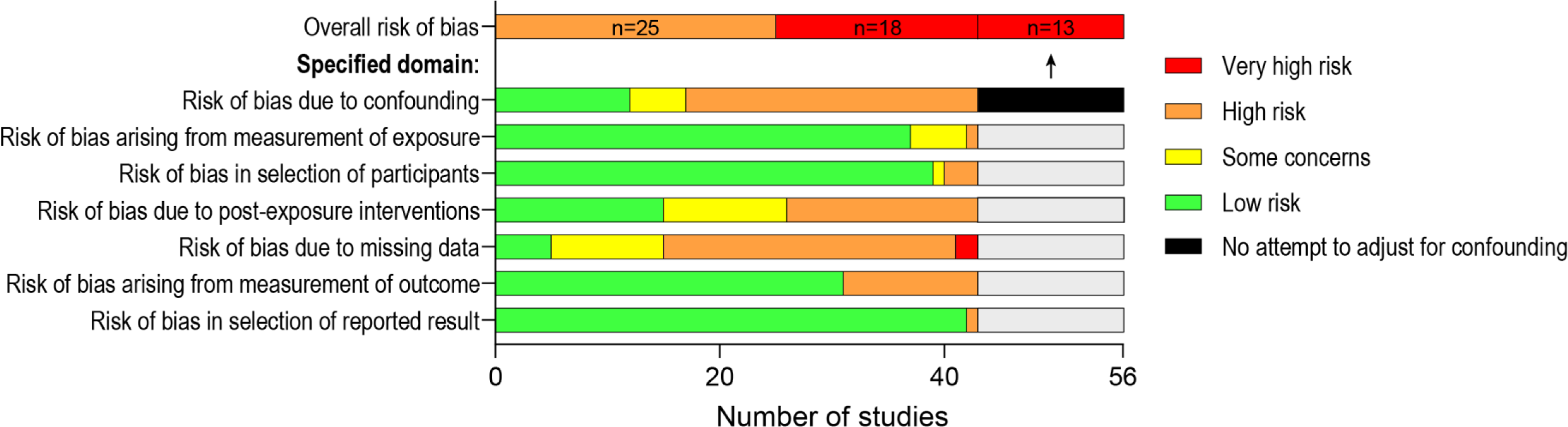

### E. Risk

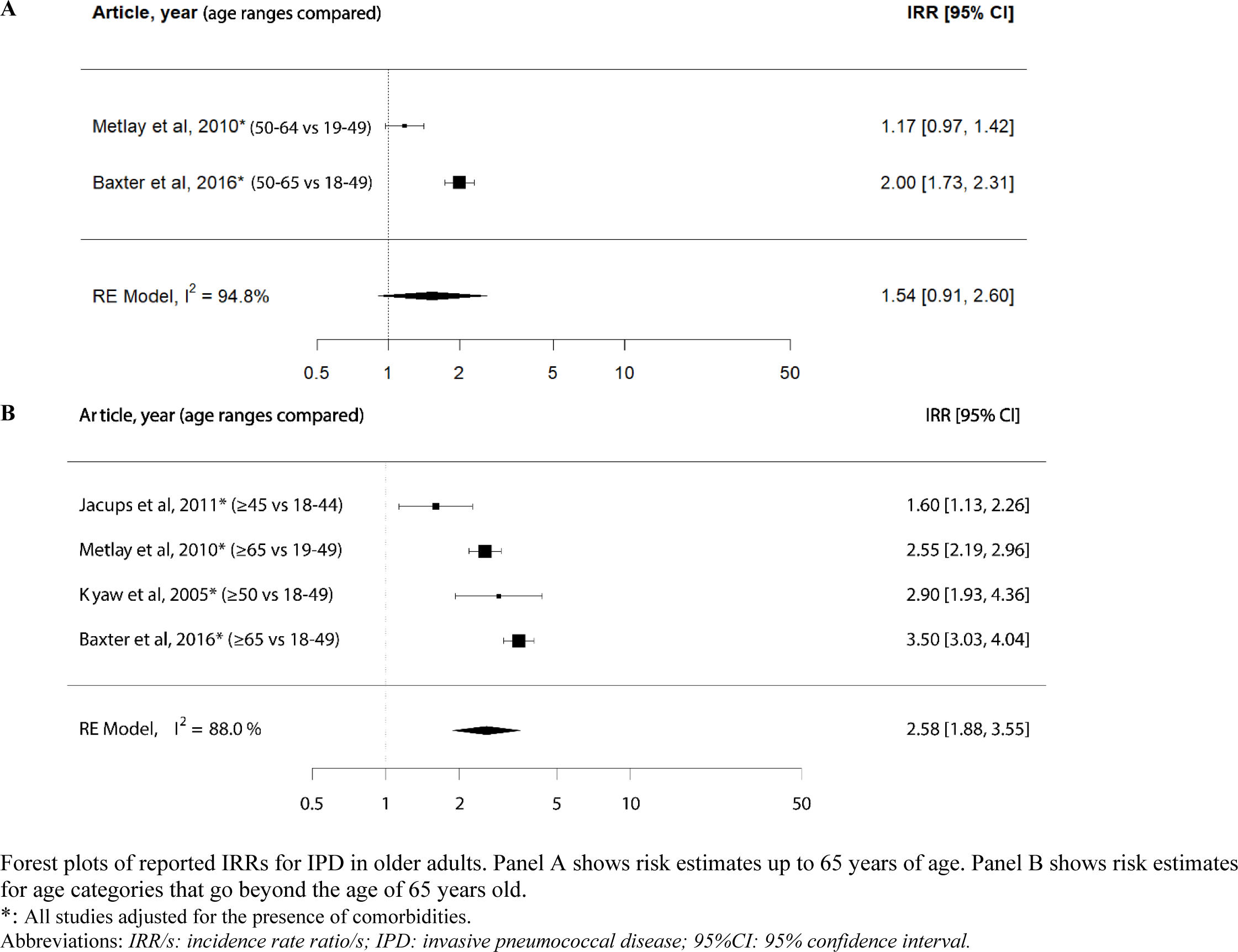

### F. Extracted data per article

Data is provided in a separate supplementary Excel file named “Risk adult IPD_Extracted data per article.xlsx“ .

### G. Pooled risk estimates for studies reporting Ors

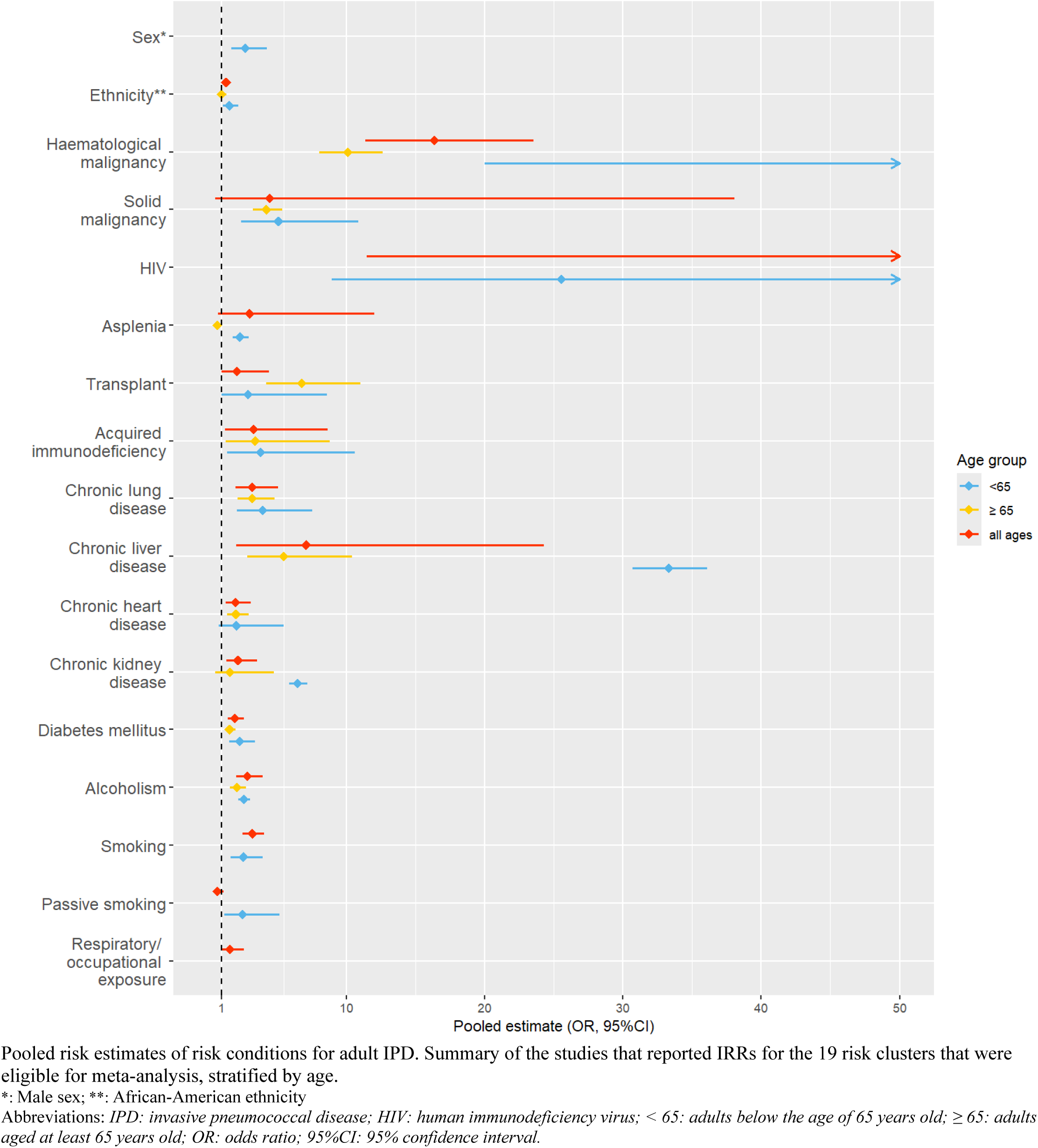

### H. Individual forest plots IRRs

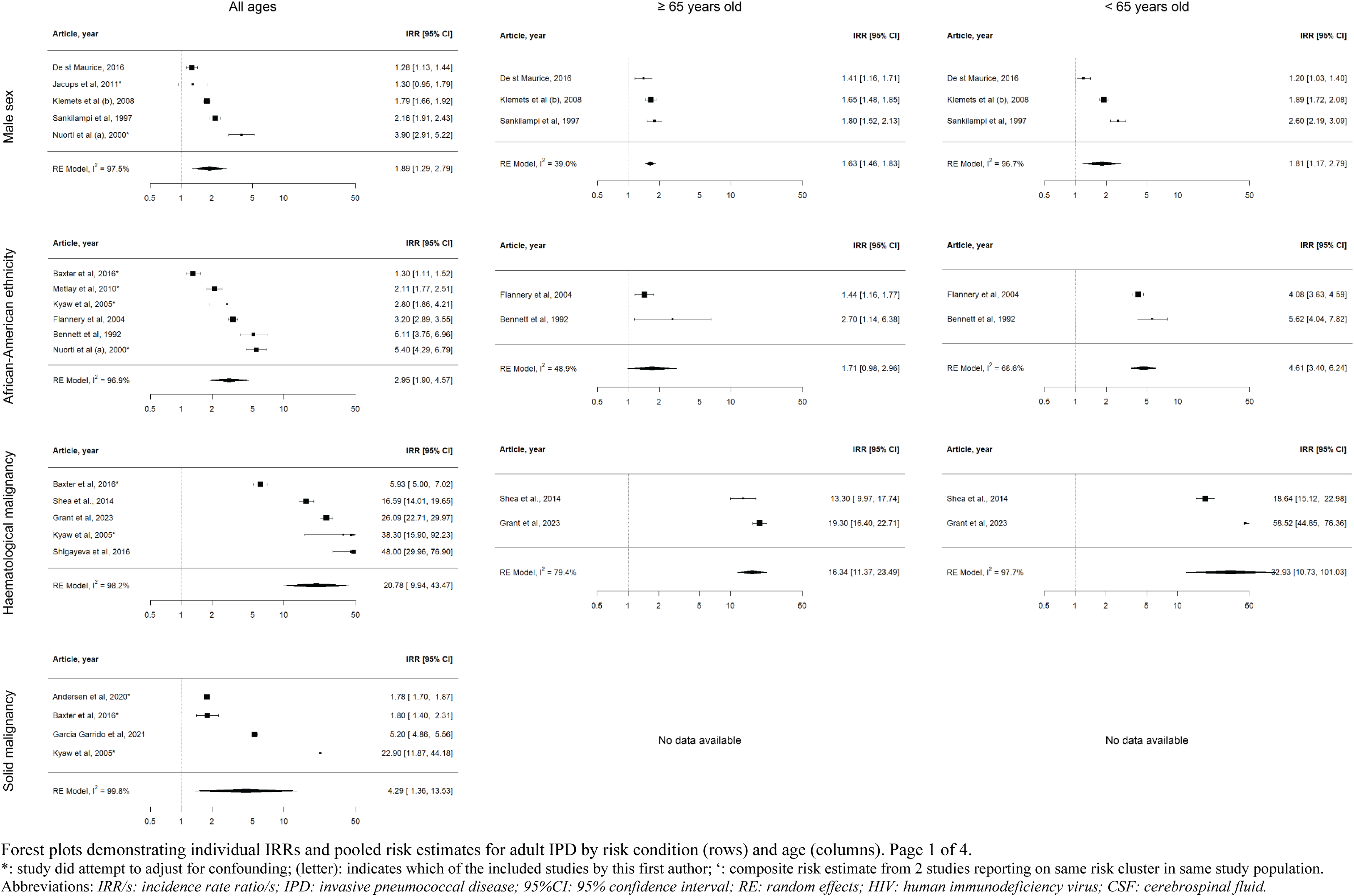
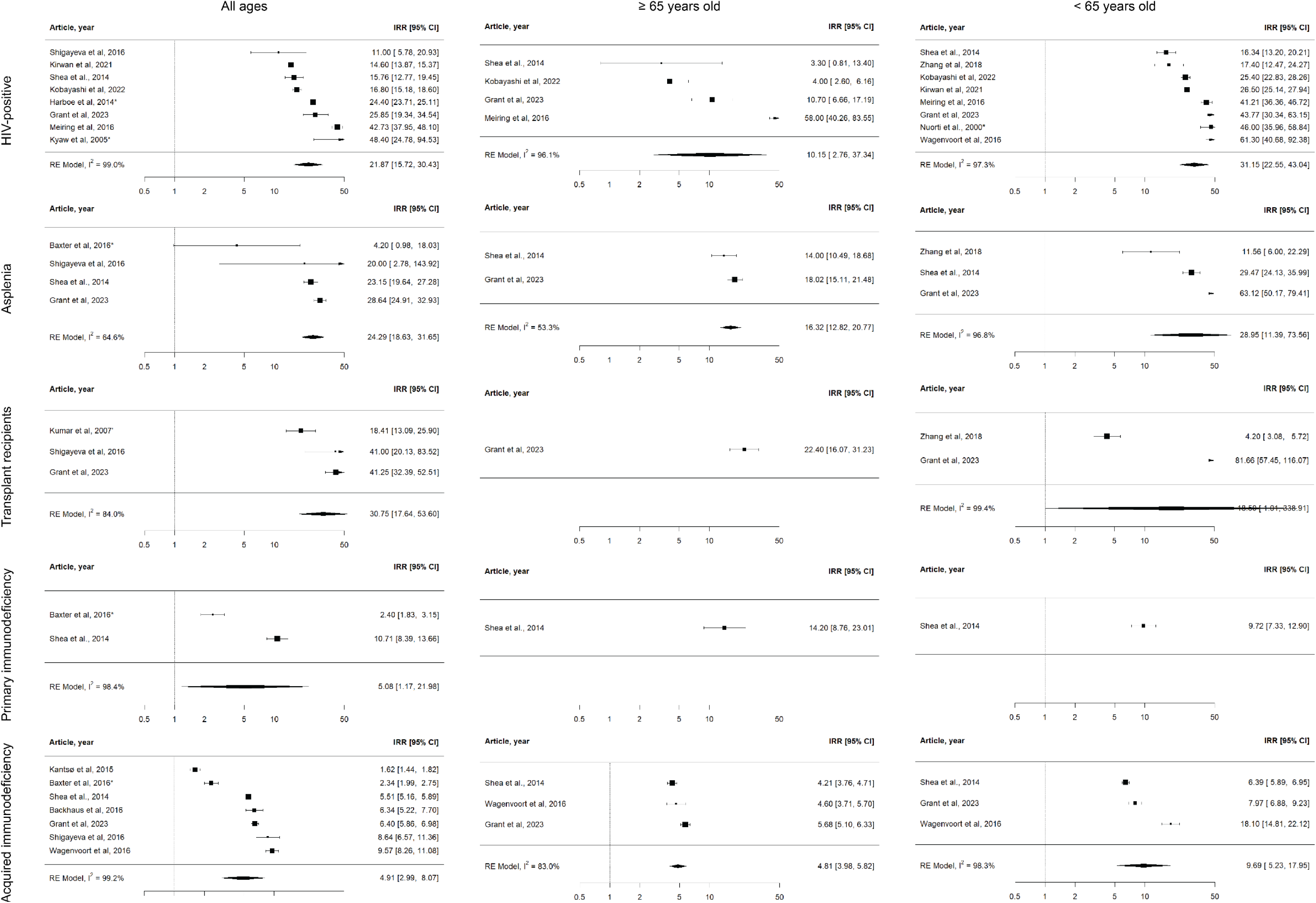
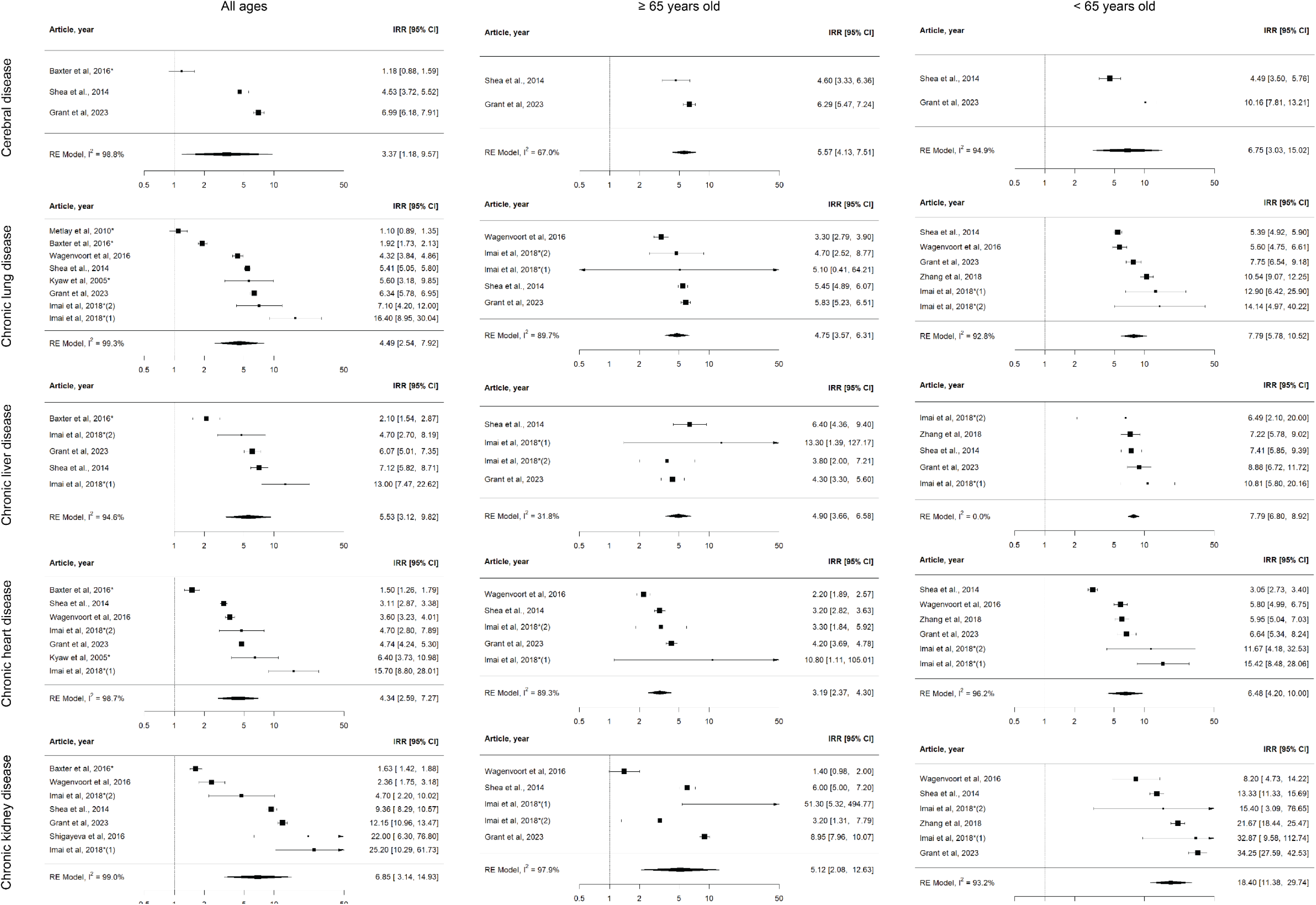
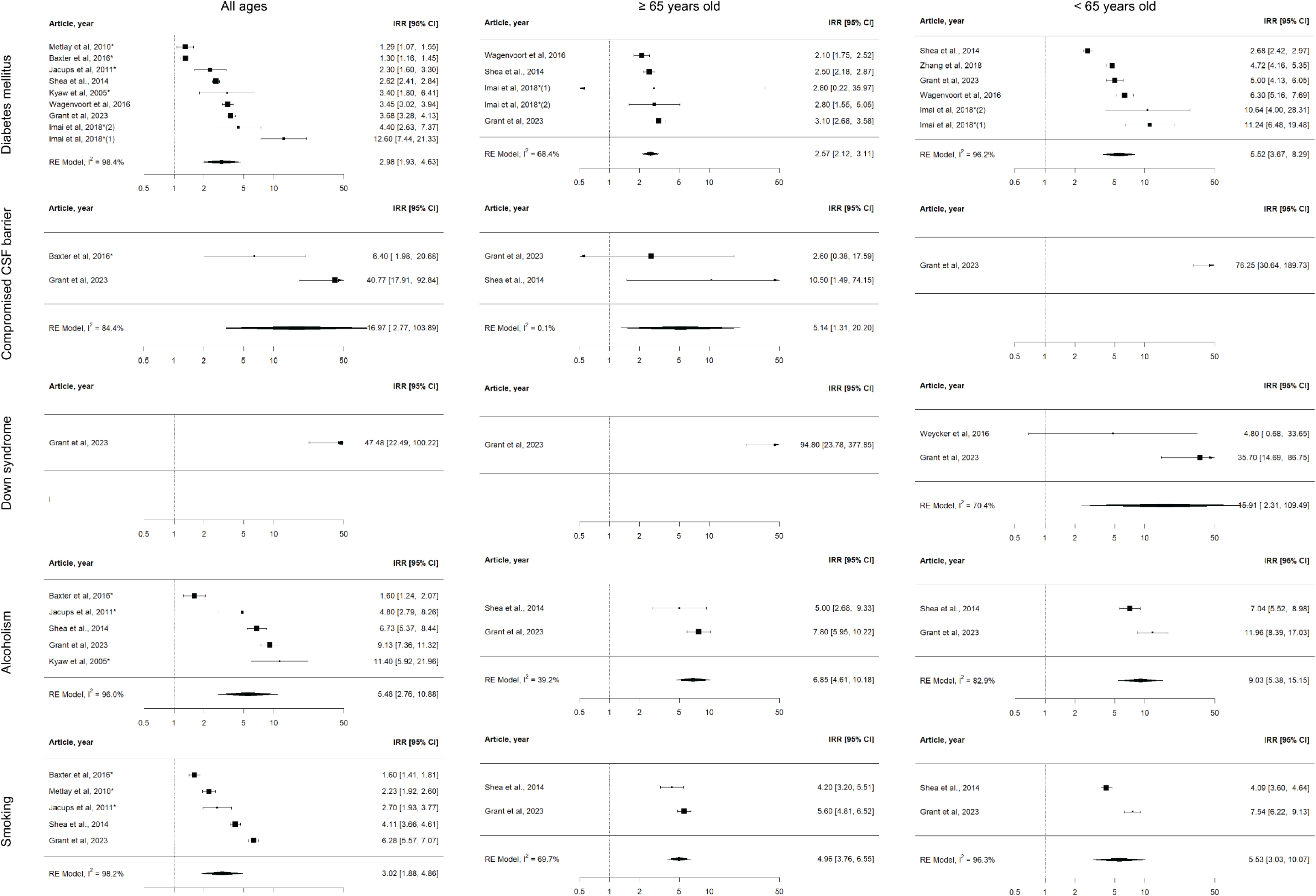

### I. Individual forest plots Ors

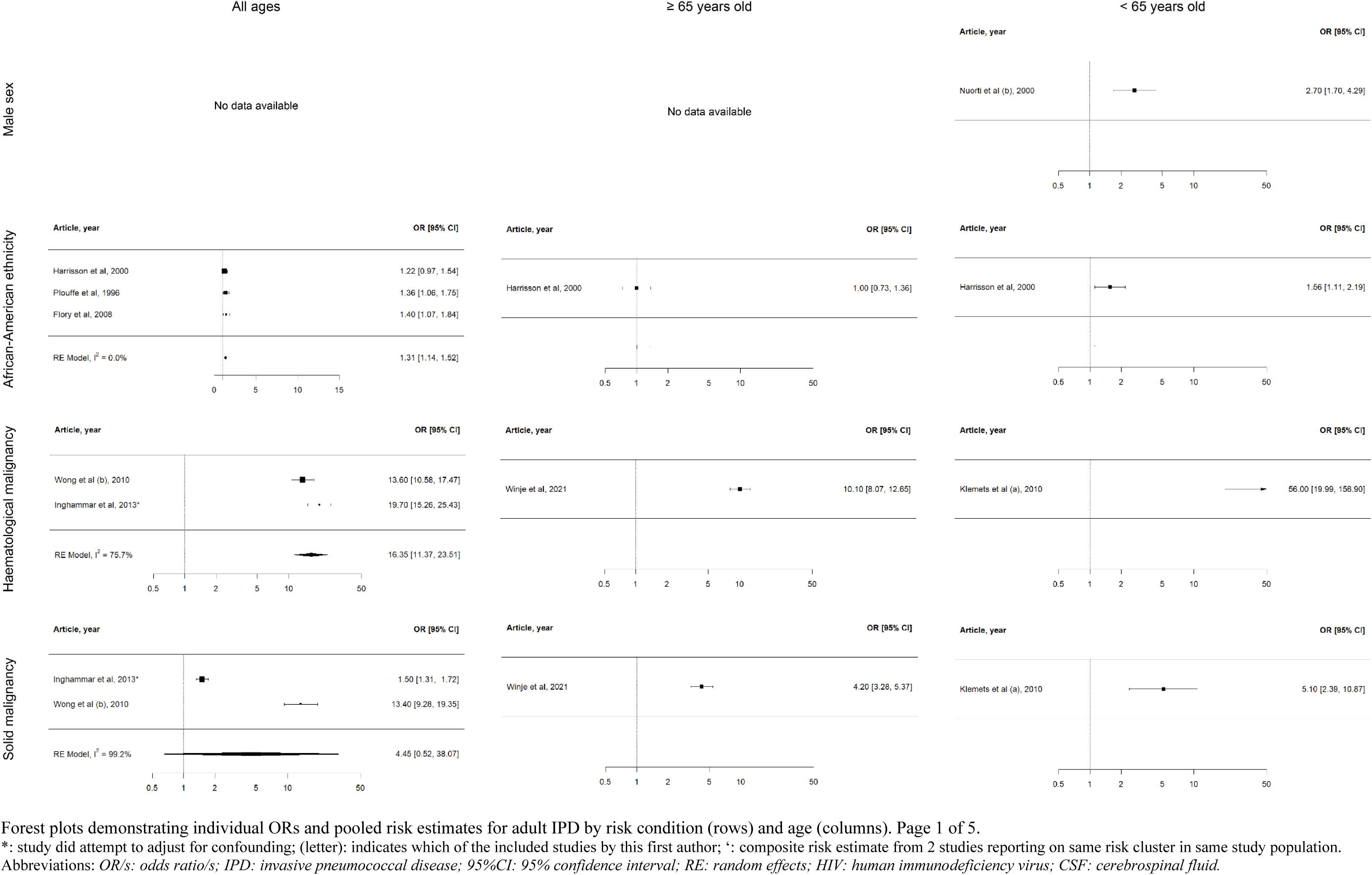
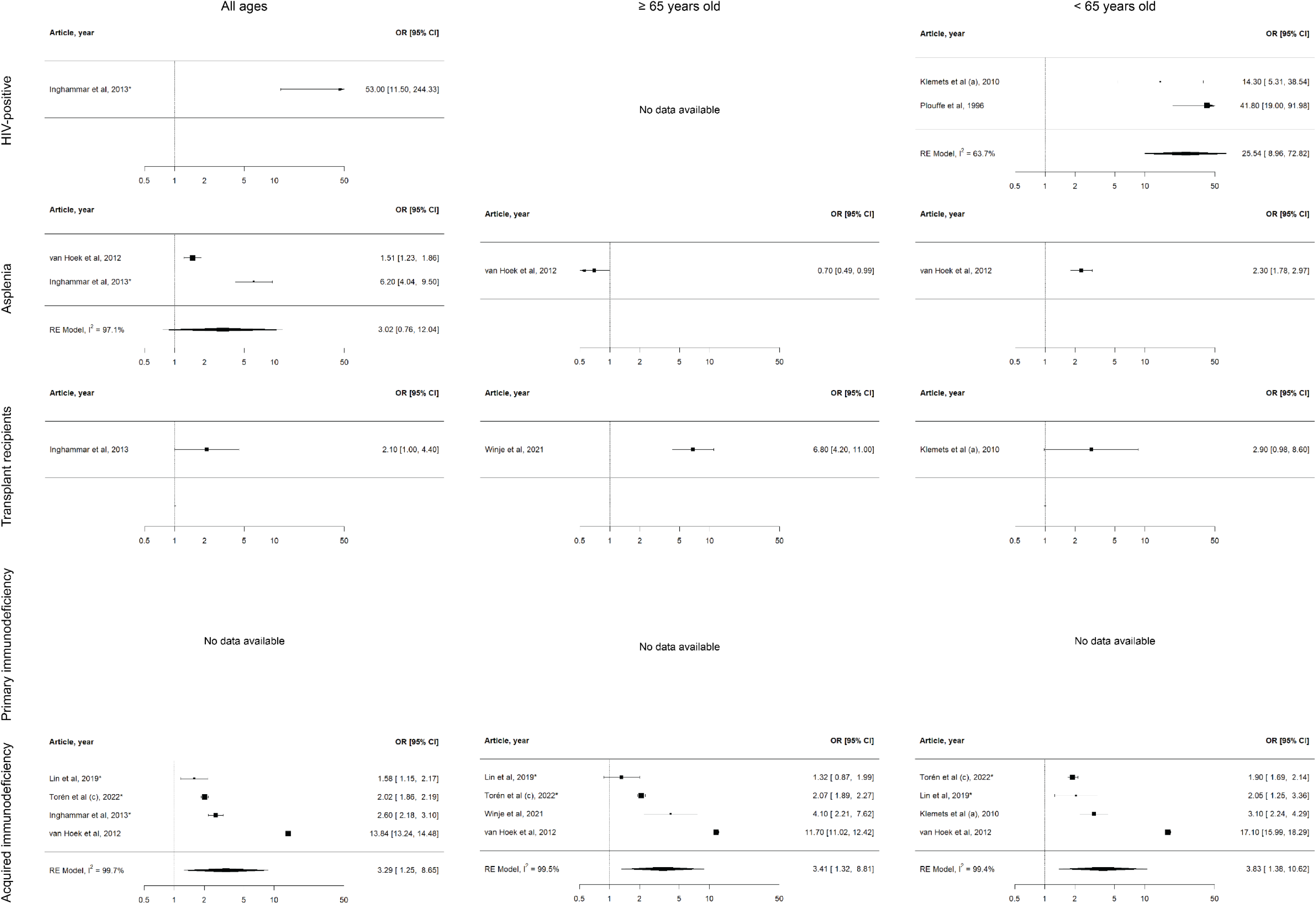
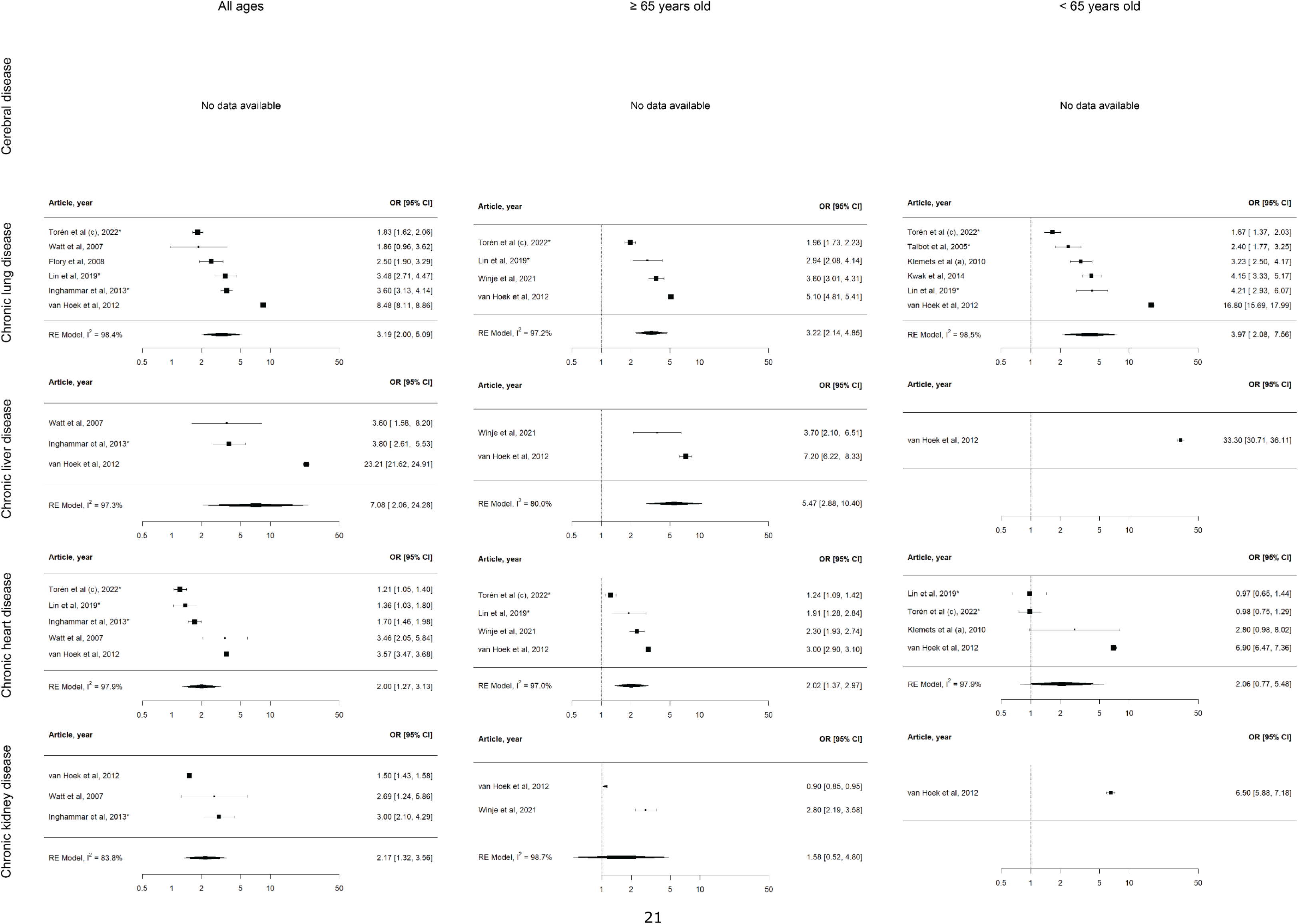
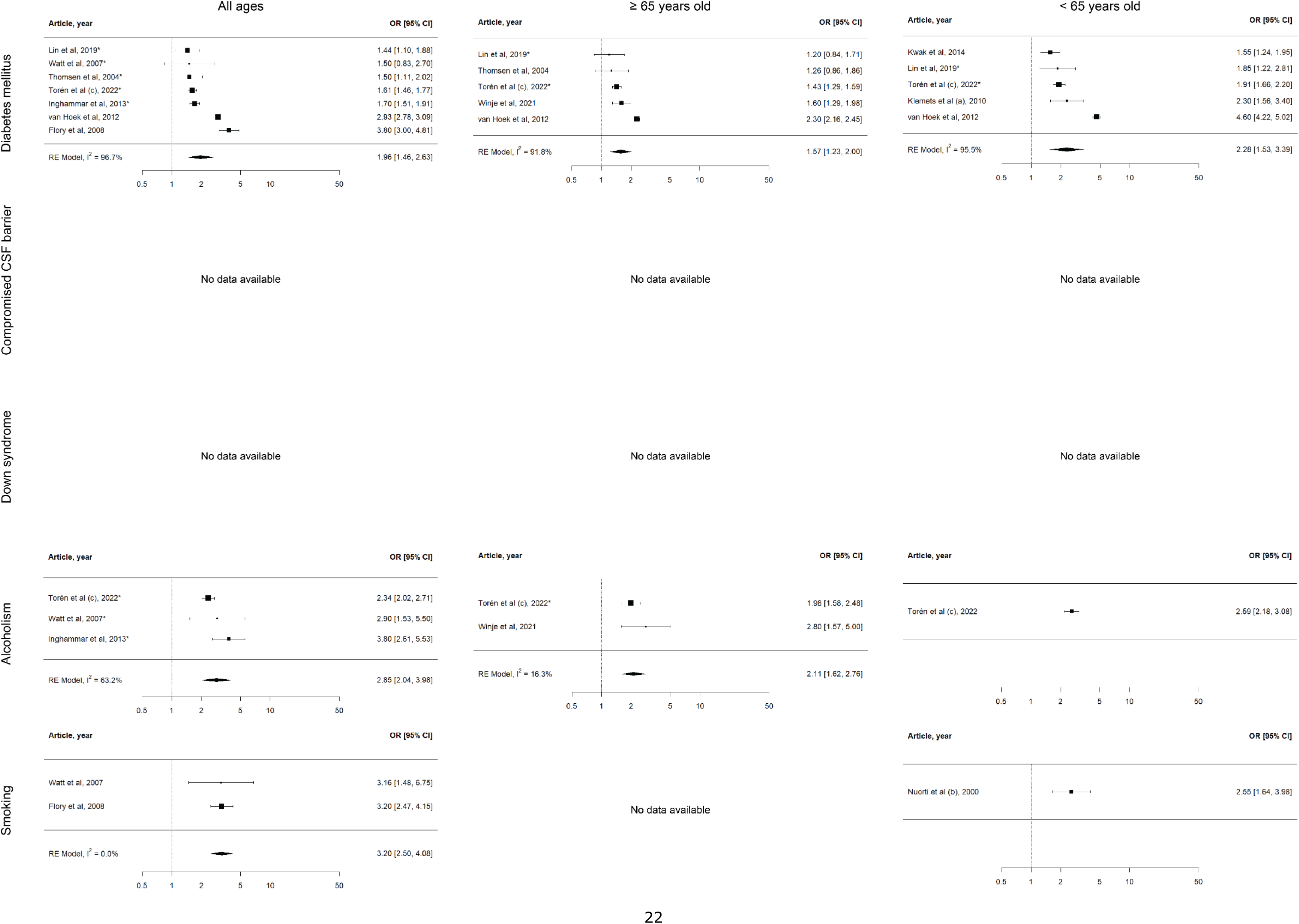
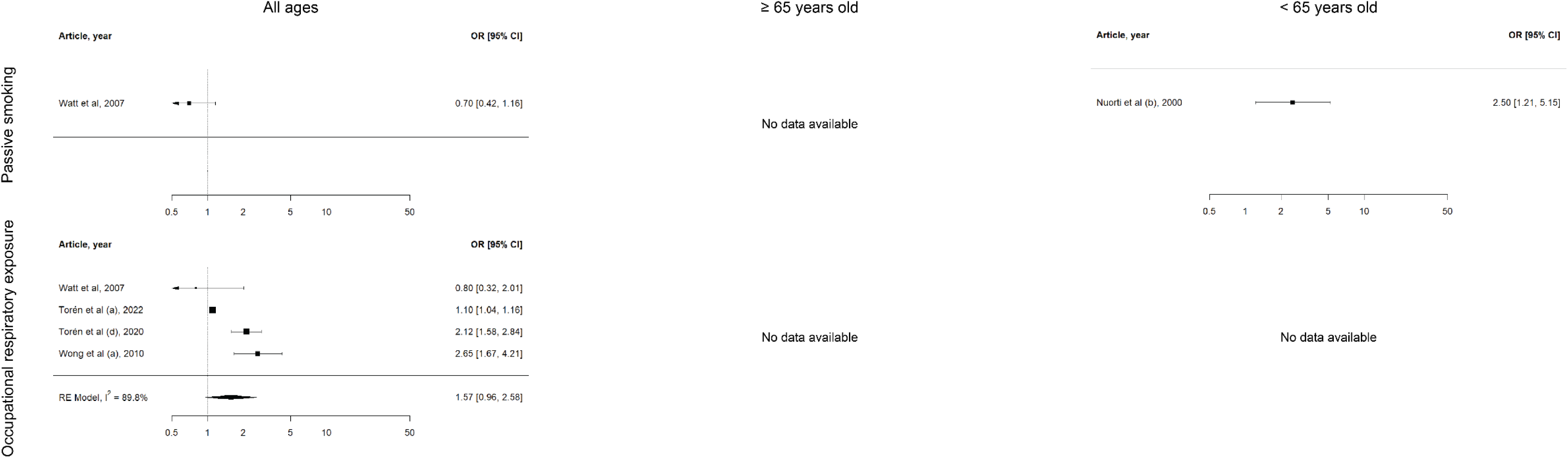

### J. Quality of evidence assessment according to GRADE system

| Risk condition | Risk of bias | Inconsistency | Indirectness | Imprecision | Publication bias | Reasons for upgrading | Quality of evidence |
| --- | --- | --- | --- | --- | --- | --- | --- |
| Male sex | Not serious | Very serious | Not serious | Not serious | Not serious | No | Very low |
| African-American ethnicity | Not serious | Very serious | Not serious | Not serious | Not serious | No | Very low |
| Haematological malignancy | Serious | Very serious | Not serious | Serious | Not serious | Yes (+1 since large effect) | Very low |
| Solid malignancy | Serious | Very serious | Not serious | Very serious | Not serious | No | Very low |
| HIV-positive | Serious | Very serious | Serious | Not serious | Not serious | Yes (+2 since large effect and lower risk if better treated) | Very low |
| Anatomic/functional asplenia | Serious | Serious | Not serious | Not serious | Not serious | Yes (+1 since large effect) | Very low |
| Transplant recipient | Serious | Serious | Not serious | Not serious | Not serious | Yes (+1 since large effect) | Very low |
| Primary immunodeficiency | Serious | Very serious | Not serious | Very serious | Not serious | No | Very low |
| Acquired immunodeficiency | Serious | Very serious | Serious | Not serious | Not serious | No | Very low |
| Cerebral disease | Serious | Very serious | Not serious | Very serious | Serious | No | Very low |
| Chronic lung disease | Serious | Very serious | Not serious | Not serious | Not serious | No | Very low |
| Chronic liver disease | Serious | Very serious | Not serious | Not serious | Not serious | No | Very low |
| Chronic heart disease | Serious | Very serious | Not serious | Not serious | Not serious | No | Very low |
| Chronic kidney disease | Serious | Very serious | Not serious | Serious | Not serious | No | Very low |
| Diabetes mellitus | Serious | Very serious | Not serious | Not serious | Not serious | No | Very low |
| Compromised CFS barrier | Serious | Serious | Not serious | Very serious | Not serious | Yes (+1 since large effect) | Very low |
| Down's syndrome | Serious | Not serious | Not serious | Very serious | Not serious | Yes (+1 since large effect) | Very low |
| Alcoholism | Serious | Very serious | Not serious | Serious | Not serious | No | Very low |
| Smoking | Serious | Very serious | Not serious | Not serious | Not serious | Yes (+1 because available data on dose-response) | Very low |

### K. Risk factors, not included in meta-analysis

| Risk factor: ethnicity (other than African-American) |  |  |  |  |  |  |  |  |
| --- | --- | --- | --- | --- | --- | --- | --- | --- |
| Article | Country | Study type | Years | IPD definition | n/N | Risk measure | Risk (95% CI) | Remarks |
| Jacups et al, 2011 | Australia | Cohort | 1987-2008 | Positive cultures | 205/114537 | aIRR | 5.9 (3.7-9.5) | Indigenous vs non-indigenous |
| Flory et al, 2008 | United States | Cohort | 2002-2004 | Positive cultures | 609/2881132 | aOR | 8.7 (3.1-24.5) | Native American |
| Flory et al, 2008 | United States | Cohort | 2002-2004 | Positive cultures | 609/2881132 | aOR | 0.4 (0.1-3.1) | Asian-American |
| Torzillo et al, 1995 | Australia | Cohort | 1985-1991 | Positive cultures | 78/31510 | RR | 20.4 (9.7-42.5) | Age 15-19. Aboriginal |
| Torzillo et al, 1995 | Australia | Cohort | 1985-1991 | Positive cultures | 78/31510 | RR | 4.0 (1.1-15.0) | Age 60+. Aboriginal |
| Metlay et al, 2010 | United states | Cohort | 2005-2008 | Positive cultures | 1376/3.7mln | aIRR | 0.70 (0.36-1.35) | Hispanic vs non-Hispanic |
| Baxter et al, 2016 | United States | Cohort | 2008-2014 | Positive cultures | 1549/15102047* | aIRR | 0.7 (0.6-0.8) | Asian vs white |
| Baxter et al, 2016 | United States | Cohort | 2008-2014 | Positive cultures | 1549/15102047* | aIRR | 1.0 (0.9-1.2) | Hispanic vs white |
| Baxter et al, 2016 | United States | Cohort | 2008-2014 | Positive cultures | 1549/15102047* | aIRR | 1.3 (0.8-2.0) | Other vs white |
| Steinberg et al, 2024 | United States | Cohort | 2011-2020 | Positive cultures | 1164/u | aIRR | 4.7 (4.2-5.2) | Native Alaska vs non-native |

| Risk factor: demography |  |  |  |  |  |  |  |  |
| --- | --- | --- | --- | --- | --- | --- | --- | --- |
| Article | Country | Study type | Years | IPD definition | n/N | Risk measure | Risk (95% CI) | Remarks |
| Feemster et al, 2013 | United States | Cohort | 2005-2008 | Positive cultures | 1416/3849071 | aIRR | 1.10 (1.00-1.19) | Density (10000 people/square mile increase) |
| Feemster et al, 2013 | United States | Cohort | 2005-2008 | Positive cultures | 1416/3849071 | aIRR | 1.07 (1.04-1.09) | % black population (10% increase) |
| Feemster et al, 2013 | United States | Cohort | 2005-2008 | Positive cultures | 1416/3849071 | aIRR | 3.49 (1.80-5.18) | % population aged < 5 yo (10% increase) |
| Feemster et al, 2013 | United States | Cohort | 2005-2008 | Positive cultures | 1416/3849071 | aIRR | 1.19 (1.00-1.38) | % population aged ≥65 yo (10% increase) |
| Nuorti et al (b), 2000 | Canada and | Case control | 1995-1996 | Positive cultures | 228/529 | OR | 3.0 (1.5-6.2) | Children in household (<6 yo in day care) |
| Nuorti et al (b), 2000 | Canada and United States | Case control | 1995-1996 | Positive cultures | 228/529 | OR | 1.0 (0.5-2.0) | Children in household (<6 yo, not in day care) |
| Metlay et al, 2010 | United States | Cohort | 2005-2008 | Positive cultures | 1376/3.7mln | aIRR | 0.47 (0.35-0.61) | 1 child aged <7y in home |
| Metlay et al, 2010 | United States | Cohort | 2005-2008 | Positive cultures | 1376/3.7mln | aIRR | 0.24 (0.18-0.33) | ≥ 2 children aged <7y in home |
| Haglund et al, 1993 | United States | Cohort | 1989-1990 | Positive cultures | 144/848273 | IRR | 13.7 (7.7-24.7) | Nursing home resident |
| Watt et al, 2007 | United States | Case control | 1999-2002 | Positive cultures | 118/417 | OR | 0.9 (0.7-1.2) | Living with a child aged ≤5 |
| Watt et al, 2007 | United States | Case control | 1999-2002 | Positive cultures | 118/417 | OR | 0.9 (0.3-3.0) | Living with a child aged ≤5 who attended day care |
| Watt et al, 2007 | United States | Case control | 1999-2002 | Positive cultures | 118/417 | OR | 0.8 (0.3-2.3) | Living with an unvaccinated child aged ≤5 |
| Watt et al, 2007 | United States | Case control | 1999-2002 | Positive cultures | 118/417 | OR | 0.8 (0.4-1.4) | Living with an unvaccinated child any age |
| Kupronis et al, 2000 | United States | Cohort | 2000-2001 | Positive cultures | 1258/2.4mln | IRR | 4.3 (3.7-5.0) | Long-term care facilities |
| Chapman et al, 2013 | United Kingdom | Cohort | 2006-2011 | Positive cultures | 1351/2148629 | aRR | 0.83 (0.7-0.99) | Age ≥16. Town vs urban (>10k) |
| Chapman et al, 2013 | United Kingdom | Cohort | 2006-2011 | Positive cultures | 1351/2148629 | aRR | 0.95 (0.75-1.22) | Age 16-64. Town vs urban (>10k) |
| Chapman et al, 2013 | United Kingdom | Cohort | 2006-2011 | Positive cultures | 1351/2148629 | aRR | 0.74 (0.57-0.95) | Age ≥65. Town vs urban (>10k) |
| Chapman et al, 2013 | United Kingdom | Cohort | 2006-2011 | Positive cultures | 1351/2148629 | aRR | 0.51 (0.37-0.71) | Age ≥16. Village vs urban (>10k) |
| Chapman et al, 2013 | United Kingdom | Cohort | 2006-2011 | Positive cultures | 1351/2148629 | aRR | 0.58 (0.36-0.91) | Age 16-64. Village vs urban (>10k) |
| Chapman et al, 2013 | United Kingdom | Cohort | 2006-2011 | Positive cultures | 1351/2148629 | aRR | 0.46 (0.29-0.74) | Age ≥65. Village vs urban (>10k) |

| Risk factor: socioeconomic factors |  |  |  |  |  |  |  |  |
| --- | --- | --- | --- | --- | --- | --- | --- | --- |
| Article | Country | Study type | Years | IPD definition | n/N | Risk measure | Risk (95% CI) | Remarks |
| Chapman et al, 2013 | United Kingdom | Cohort | 2006-2011 | Positive cultures | 1351/2148629 | aRR | 1.34 (1.08-1.64) | Age ≥16. Deprivation quintile 2 |
| Chapman et al, 2013 | United Kingdom | Cohort | 2006-2011 | Positive cultures | 1351/2148629 | aRR | 1.40 (1.02-1.93) | Age 16-64. Deprivation quintile 2 |
| Chapman et al, 2013 | United Kingdom | Cohort | 2006-2011 | Positive cultures | 1351/2148629 | aRR | 1.27 (0.97-1.68) | Age ≥65. Deprivation quintile 2 |
| Chapman et al, 2013 | United Kingdom | Cohort | 2006-2011 | Positive cultures | 1351/2148629 | aRR | 1.50 (1.22-1.85) | Age ≥16. Deprivation quintile 3 |
| Chapman et al, 2013 | United Kingdom | Cohort | 2006-2011 | Positive cultures | 1351/2148629 | aRR | 1.75 (1.29-2.38) | Age 16-64. Deprivation quintile 3 |
| Chapman et al, 2013 | United Kingdom | Cohort | 2006-2011 | Positive cultures | 1351/2148629 | aRR | 1.31 (0.99-1.73) | Age ≥65. Deprivation quintile 3 |
| Chapman et al, 2013 | United Kingdom | Cohort | 2006-2011 | Positive cultures | 1351/2148629 | aRR | 1.86 (1.53-2.28) | Age ≥16. Deprivation quintile 4 |
| Chapman et al, 2013 | United Kingdom | Cohort | 2006-2011 | Positive cultures | 1351/2148629 | aRR | 2.03 (1.50-2.75) | Age 16-64. Deprivation quintile 4 |
| Chapman et al, 2013 | United Kingdom | Cohort | 2006-2011 | Positive cultures | 1351/2148629 | aRR | 1.73 (1.32-2.26) | Age ≥65. Deprivation quintile 4 |
| Chapman et al, 2013 | United Kingdom | Cohort | 2006-2011 | Positive cultures | 1351/2148629 | aRR | 2.44 (2.01-2.96) | Age ≥16. Deprivation quintile 5 |
| Chapman et al, 2013 | United Kingdom | Cohort | 2006-2011 | Positive cultures | 1351/2148629 | aRR | 3.11 (2.35-4.14) | Age 16-64. Deprivation quintile 5 |
| Chapman et al, 2013 | United Kingdom | Cohort | 2006-2011 | Positive cultures | 1351/2148629 | aRR | 1.89 (1.44-2.47) | Age ≥65. Deprivation quintile 5 |
| Watt et al, 2007 | United States | Case control | 1999-2002 | Positive cultures | 118/417 | aOR | 2.6 (1.2-5.5) | Unemployment |
| Nuorti et al (b), 2000 | Canada and US | Case control | 1995-1996 | Positive cultures | 228/529 | OR | 2.8 (1.3-5.9) | Less than high school (compared to college) |
| Nuorti et al (b), 2000 | Canada and US | Case control | 1995-1996 | Positive cultures | 228/529 | OR | 2.0 (1.2-3.4) | High school versus college |
| Harrison et al, 2000 | United States | Cohort | 1995-1996 | Positive cultures | 1051/5mln | aRR | 2.1 (0.8-5.2) | Age 20-29. Low vs high income |
| Harrison et al, 2000 | United States | Cohort | 1995-1996 | Positive cultures | 1051/5mln | aRR | 6.7 (3.9-11.7) | Age 30-44. Low vs high income |
| Harrison et al, 2000 | United States | Cohort | 1995-1996 | Positive cultures | 1051/5mln | aRR | 3.3 (2.2-4.9) | Age 45-64. Low vs high income |
| Harrison et al, 2000 | United States | Cohort | 1995-1996 | Positive cultures | 1051/5mln | aRR | 1.6 (1.3-2.1) | Age 65+. Low vs high income |
| Inghammar et al, 2013 | Sweden | Case control | 1990-2008 | Positive cultures | 4085/44438 | OR | 1.9 (1.2-2.9) | Secondary education (vs primary) |
| Inghammar et al, 2013 | Sweden | Case control | 1990-2008 | Positive cultures | 4085/44438 | aOR | 1.8 (1.1-2.38) | Upper secondary education (vs primary) |
| Inghammar et al, 2013 | Sweden | Case control | 1990-2008 | Positive cultures | 4085/44438 | aOR | 1.5 (1.0-1.0) | College/university (vs primary) \| Adjusted OR |
| Flory et al, 2008 | United States | Cohort | 2002-2004 | Positive cultures | 609/2881132 | aOR | 11.5 (7.4-18.1) | Annual income < \$6000 (compared to > 50000) |
| Flory et al, 2008 | United States | Cohort | 2002-2004 | Positive cultures | 609/2881132 | aOR | 6.0 (3.7-9.6) | Annual income \$6000-11999 (compared to > \$50000) |
| Flory et al, 2008 | United States | Cohort | 2002-2004 | Positive cultures | 609/2881132 | aOR | 3.1 (1.7-5.7) | Annual income \$12000-24999 (compared to > \$50000) |
| Flory et al, 2008 | United States | Cohort | 2002-2004 | Positive cultures | 609/2881132 | aOR | 2.1 (1.4-3.2) | Annual income \$25000-49999 (compared to > \$50000) |
| Flory et al, 2008 | United States | Cohort | 2002-2004 | Positive cultures | 609/2881132 | aOR | 0.7 (0.5-1.1) | No high school diploma (vs college graduate) |
| Flory et al, 2008 | United States | Cohort | 2002-2004 | Positive cultures | 609/2881132 | aOR | 0.6 (0.4-0.8) | High school diploma (vs college graduate) |
| Flory et al, 2008 | United States | Cohort | 2002-2004 | Positive cultures | 609/2881132 | aOR | 0.7 (0.5-1.1) | Some college (vs college graduate) |
| Steinberg et al, 2024 | United States | Cohort | 2011-2020 | Positive cultures | 1164/u | aIRR | 72 (59-89) | Homelessness |

| Risk factor: weight |  |  |  |  |  |  |  |  |
| --- | --- | --- | --- | --- | --- | --- | --- | --- |
| Article | Country | Study type | Years | IPD definition | n/N | Risk measure | Risk (95% CI) | Remarks |
| Watt et al, 2007 | United States | Case control | 1999-2002 | Positive cultures | 118/417 | aOR | 3.2 (0.98-10.6) | BMI <5th percentile |
| Watt et al, 2007 | United States | Case control | 1999-2002 | Positive cultures | 118/417 | aOR | 2.8 (0.97-8.0) | BMI >95th percentile |
| Grant et al, 2023 | United States | Cohort | 2016-2019 | ICD codes | u/38618534* | IRR | 4.0 (2.8-5.9) | Age 18-49. Obesity |
| Grant et al, 2023 | United States | Cohort | 2016-2019 | ICD codes | u/38618534* | IRR | 4.2 (3.2-5.4) | Age 50-64. Obesity |
| Grant et al, 2023 | United States | Cohort | 2016-2019 | ICD codes | u/38618534* | IRR | 2.8 (2.4-3.4) | Age ≥65. Obesity |

| Risk factor: inhalation corticosteroid use |  |  |  |  |  |  |  |  |
| --- | --- | --- | --- | --- | --- | --- | --- | --- |
| Article | Country | Study type | Years | IPD definition | n/N | Risk measure | Risk (95% CI) | Remarks |
| Torén et al (c), 2022 | Sweden | Case control | 2007-2018 | Positive cultures | 6870/48090 | aOR | 2.33 (2.06-2.62) | Current use ICS |
| Torén et al (c), 2022 | Sweden | Case control | 2007-2018 | Positive cultures | 6870/48090 | aOR | 1.64 (1.41-1.89) | Past use ICS |

| Risk factor: medication other than immunosuppressive medication |  |  |  |  |  |  |  |  |
| --- | --- | --- | --- | --- | --- | --- | --- | --- |
| Article | Country | Study type | Years | IPD definition | n/N | Risk measure | Risk (95% CI) | Remarks |
| Wiese et al, 2018 | United States | Case control | 1995-2014 | Positive cultures | 1233/25632 | aOR | 1,62 (1.36-1,92) | Current opioid use |
| Torén et al (c), 2022 | Sweden | Case control | 2007-2018 | Positive cultures | 6870/48090 | aOR | 3.90 (2.48-6.13) | Age 20-80. Use of immunostimulating drugs ≤12 mo |
| Torén et al (c), 2022 | Sweden | Case control | 2007-2018 | Positive cultures | 6870/48090 | aOR | 3.40 (1.96-5.92) | Age 20-64. Use of immunostimulating drugs ≤12 mo |
| Torén et al (c), 2022 | Sweden | Case control | 2007-2018 | Positive cultures | 6870/48090 | aOR | 5.10 (2.56-10.15) | Age 65-80. Use of immunostimulating drugs ≤12 mo |
| Torén et al (c), 2022 | Sweden | Case control | 2007-2018 | Positive cultures | 6870/48090 | aOR | 1.26 (1.17-1.37) | Age 20-80. Use of PPI or H2 blocker ≤12 mo |
| Torén et al (c), 2022 | Sweden | Case control | 2007-2018 | Positive cultures | 6870/48090 | aOR | 1.33 (1.19-1.49) | Age 20-64. Use of PPI or H2 blocker ≤12 mo |
| Torén et al (c), 2022 | Sweden | Case control | 2007-2018 | Positive cultures | 6870/48090 | aOR | 1.23 (1.12-1.35) | Age 65-80. Use of PPI or H2 blocker ≤12 mo |
| Torén et al (c), 2022 | Sweden | Case control | 2007-2018 | Positive cultures | 6870/48090 | aOR | 1.08 (1.00-1.16) | Age 20-80. Use of antihypertensive medication ≤12 mo |
| Torén et al (c), 2022 | Sweden | Case control | 2007-2018 | Positive cultures | 6870/48090 | aOR | 1.07 (0.97-1.19) | Age 20-64. Use of antihypertensive medication ≤12 mo |
| Torén et al (c), 2022 | Sweden | Case control | 2007-2018 | Positive cultures | 6870/48090 | aOR | 1.00 (0.92-1.09) | Age 65-80. Use of antihypertensive medication ≤12 mo |
| Torén et al (c), 2022 | Sweden | Case control | 2007-2018 | Positive cultures | 6870/48090 | aOR | 1.35 (1.17-1.56) | Age 20-80. Use of antibacterial drugs ≤30d |
| Torén et al (c), 2022 | Sweden | Case control | 2007-2018 | Positive cultures | 6870/48090 | aOR | 1.38 (1.14-1.68) | Age 20-64. Use of antibacterial drugs ≤30d |
| Torén et al (c), 2022 | Sweden | Case control | 2007-2018 | Positive cultures | 6870/48090 | aOR | 1.29 (1.09-1.53) | Age 65-80. Use of antibacterial drugs ≤30d |

| Risk factor: coeliac disease |  |  |  |  |  |  |  |  |
| --- | --- | --- | --- | --- | --- | --- | --- | --- |
| Article | Country | Study type | Years | IPD definition | n/N | Risk measure | Risk (95% CI) | Remarks |
| Röckert Tjernberg et al, 2017 | Sweden | Case control | 1969-2009 | Positive cultures | 207/173269 | HR | 1.18 (0.49-2.85) | Age 20-39 |
| Röckert Tjernberg et al, 2017 | Sweden | Case control | 1969-2009 | Positive cultures | 207/173269 | HR | 1.81 (1.11-2.96) | Age 40-59 |
| Röckert Tjernberg et al, 2017 | Sweden | Case control | 1969-2009 | Positive cultures | 207/173269 | HR | 1.08 (0.59-1.98) | Age ≥60 |

| Risk factor: short gestation/low birthweight |
| --- |

| Article | Country | Study type | Years | IPD definition | n/N | Risk measure | Risk (95% CI) | Remarks |
| --- | --- | --- | --- | --- | --- | --- | --- | --- |
| Weycker et al, 2016 | United States | Cohort | 2007-2010 | ICD codes | u/91620399* | IRR | 2.1 (0.5-8.6) | Age 18-64 |

Risk factor: occupational exposures other than respiratory
| Article | Country | Study type | Years | IPD definition | n/N | Risk measure | Risk (95% CI) | Remarks |
| --- | --- | --- | --- | --- | --- | --- | --- | --- |
| Wong et al (a), 2010 | Canada | Cohort | 2000-2004 | Positive cultures | 863/u | OR | 1.2 (0.6-2.4) | Day care workers |
| Wong et al (a), 2010 | Canada | Cohort | 2000-2004 | Positive cultures | 863/u | OR | 0.35 (0.17-0.73) | Farmer |
| Wong et al (a), 2010 | Canada | Cohort | 2000-2004 | Positive cultures | 863/u | OR | 1.41 (0.67-3.01) | Electrician |
| Torén et al (b), 2022 | Sweden | Case control | 2006-2019 | Positive cultures | 3968/21666 | aOR | 1.11 (1.01-1.22) | ≤30 workers in close proximity per day |
| Torén et al (b), 2022 | Sweden | Case control | 2006-2019 | Positive cultures | 3968/21666 | aOR | 1.14 (1.02-1.26) | >30 workers in close proximity per day |
| Torén et al (b), 2022 | Sweden | Case control | 2006-2019 | Positive cultures | 3968/21666 | aOR | 1.11 (1.01-1.21) | Contacts with co-workers/general public |
| Torén et al (b), 2022 | Sweden | Case control | 2006-2019 | Positive cultures | 3968/21666 | aOR | 1.13 (1.00-1.27) | Regular contacts with infected patients |
| Torén et al (b), 2022 | Sweden | Case control | 2006-2019 | Positive cultures | 3968/21666 | aOR | 1.09 (0.99-1.20) | Sharing material/surfaces with co-workers/general public |
| Torén et al (b), 2022 | Sweden | Case control | 2006-2019 | Positive cultures | 3968/21666 | aOR | 1.13 (1.02-1.24) | Frequently sharing material/surfaces with general public |
| Torén et al (b), 2022 | Sweden | Case control | 2006-2019 | Positive cultures | 3968/21666 | aOR | 1.19 (1.04-1.38) | Working mostly outside or partly inside |
| Torén et al (b), 2022 | Sweden | Case control | 2006-2019 | Positive cultures | 3968/21666 | aOR | 1.10 (1.01-1.20) | Working mostly inside |
| Torén et al (b), 2022 | Sweden | Case control | 2006-2019 | Positive cultures | 3968/21666 | aOR | 1.11 (1.01-1.21) | Social distancing: not always |
| Torén et al (b), 2022 | Sweden | Case control | 2006-2019 | Positive cultures | 3968/21666 | aOR | 1.11 (1.00-1.24) | Social distancing: never |

Risk factor: prior infection
| Article | Country | Study type | Years | IPD definition | n/N | Risk measure | Risk (95% CI) | Remarks |
| --- | --- | --- | --- | --- | --- | --- | --- | --- |
| Torén et al (c), 2022 | Sweden | Case control | 2007-2018 | Positive cultures | 6870/48090 | aOR | 5.53 (4.00-7.62) | Age 20-80. Bacterial pneumonia |
| Torén et al (c), 2022 | Sweden | Case control | 2007-2018 | Positive cultures | 6870/48090 | aOR | 3.51 (3.34-9.08) | Age 20-64. Bacterial pneumonia |
| Torén et al (c), 2022 | Sweden | Case control | 2007-2018 | Positive cultures | 6870/48090 | aOR | 4.28 (3.11-5.89) | Age 65-80. Bacterial pneumonia |
| Torén et al (c), 2022 | Sweden | Case control | 2007-2018 | Positive cultures | 6870/48090 | aOR | 2.89 (1.33-6.27) | Age 20-80. Influenza pneumonia |
| Torén et al (c), 2022 | Sweden | Case control | 2007-2018 | Positive cultures | 6870/48090 | aOR | 2.65 (0.72-9.80) | Age 20-64. Influenza pneumonia |

Risk factor: obstructive sleep apnoea
| Article | Country | Study type | Years | IPD definition | n/N | Risk measure | Risk (95% CI) | Remarks |
| --- | --- | --- | --- | --- | --- | --- | --- | --- |
| Grant et al, 2023 | United States | Cohort | 2016-2019 | ICD codes | u/38618534* | IRR | 5.7 (3.5-9.3) | Age 18-49. |
| Grant et al, 2023 | United States | Cohort | 2016-2019 | ICD codes | u/38618534* | IRR | 4.2 (3.1-5.6) | age 50-64. |
| Grant et al, 2023 | United States | Cohort | 2016-2019 | ICD codes | u/38618534* | IRR | 3.4 (2.8-4.2) | age ≥65. |

| Risk factor: metabolic disorders |  |  |  |  |  |  |  |  |
| --- | --- | --- | --- | --- | --- | --- | --- | --- |
| Article | Country | Study type | Years | IPD definition | n/N | Risk measure | Risk (95% CI) | Remarks |
| Grant et al, 2023 | United States | Cohort | 2016-2019 | ICD codes | u/38618534* | IRR | 2.3 (0.3-16.7) | Age 18-49. Unclear how defined |
| Grant et al, 2023 | United States | Cohort | 2016-2019 | ICD codes | u/38618534* | IRR | 3.2 (1.1-9.2) | age 50-64. Unclear how defined |
| Grant et al, 2023 | United States | Cohort | 2016-2019 | ICD codes | u/38618534* | IRR | 2.7 (1.7-4.3) | age $\geq 65$. Unclear how defined |

Positive cultures are defined as positive cultures from a normally sterile site. *n*=number of IPD cases, *N* = total number of participants, *u* =unknown, \*=person-years.

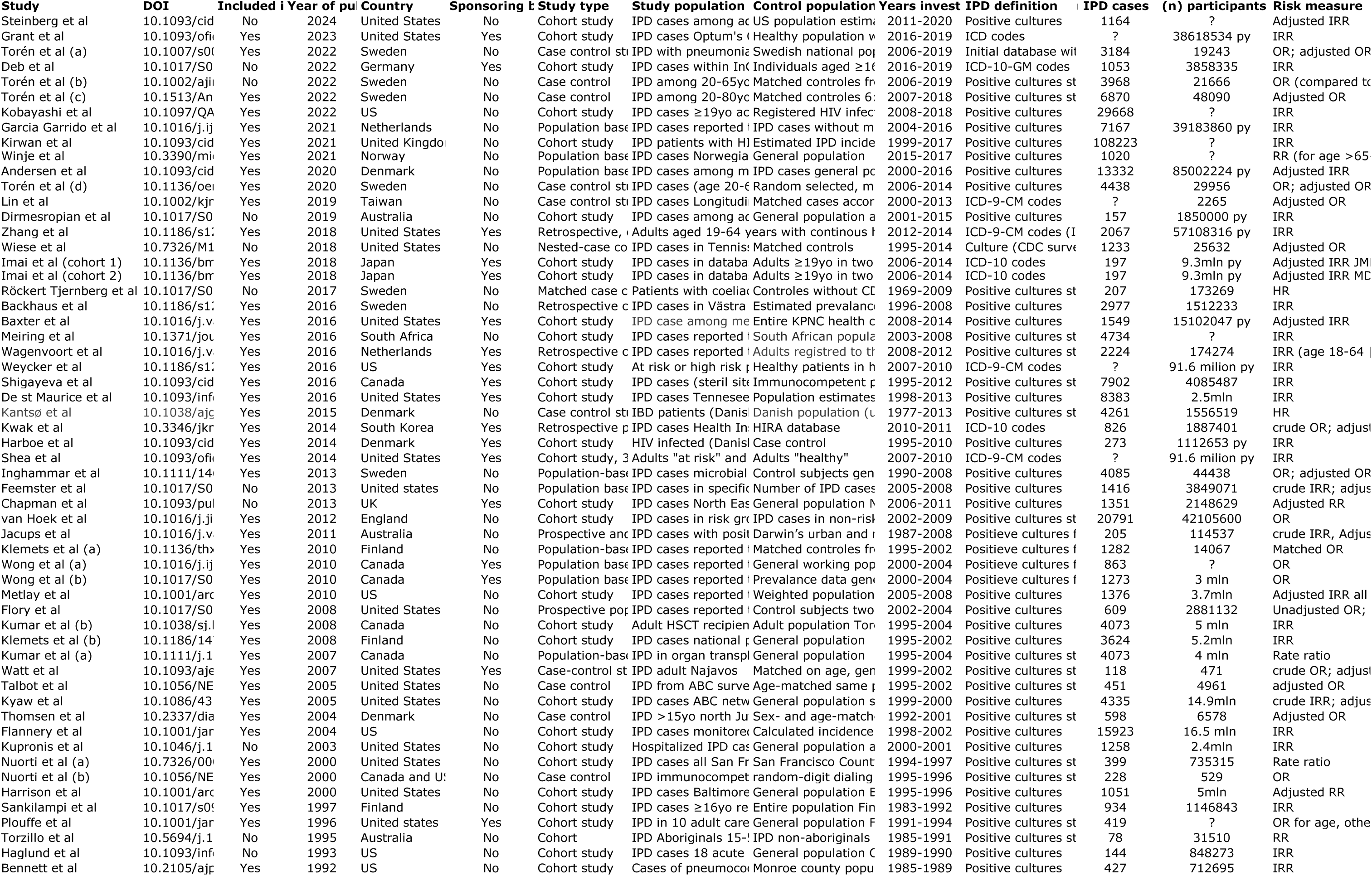
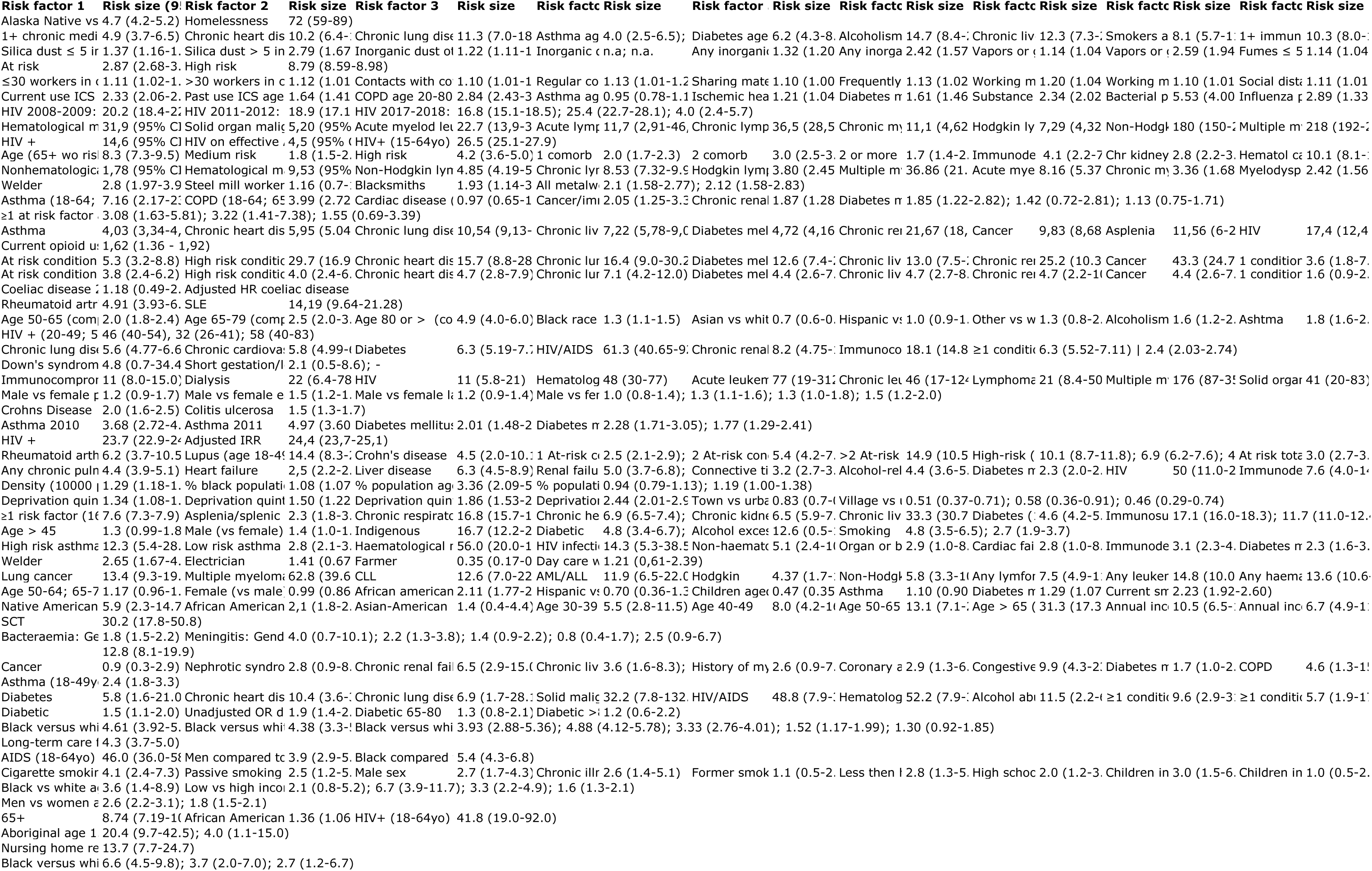
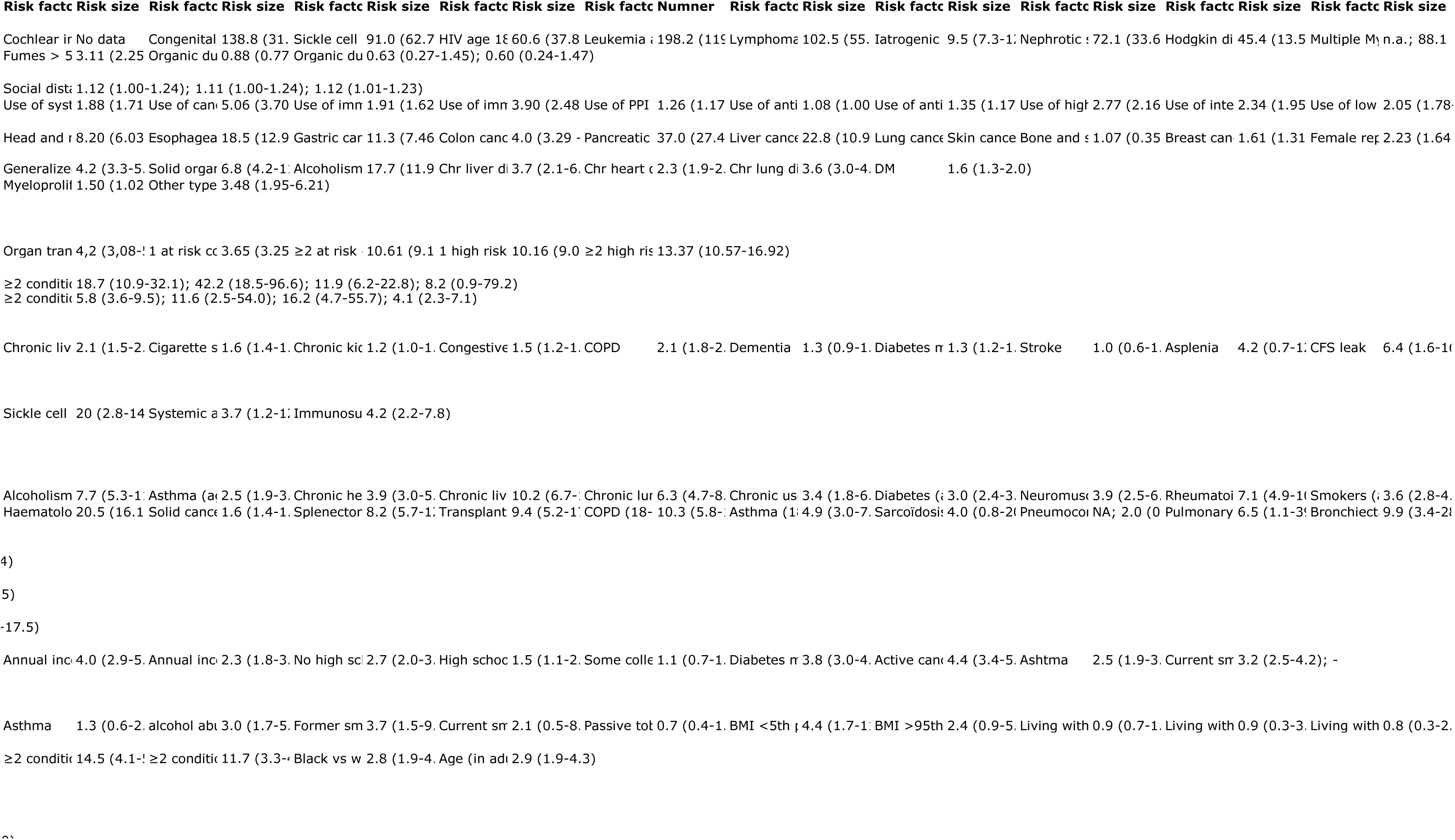
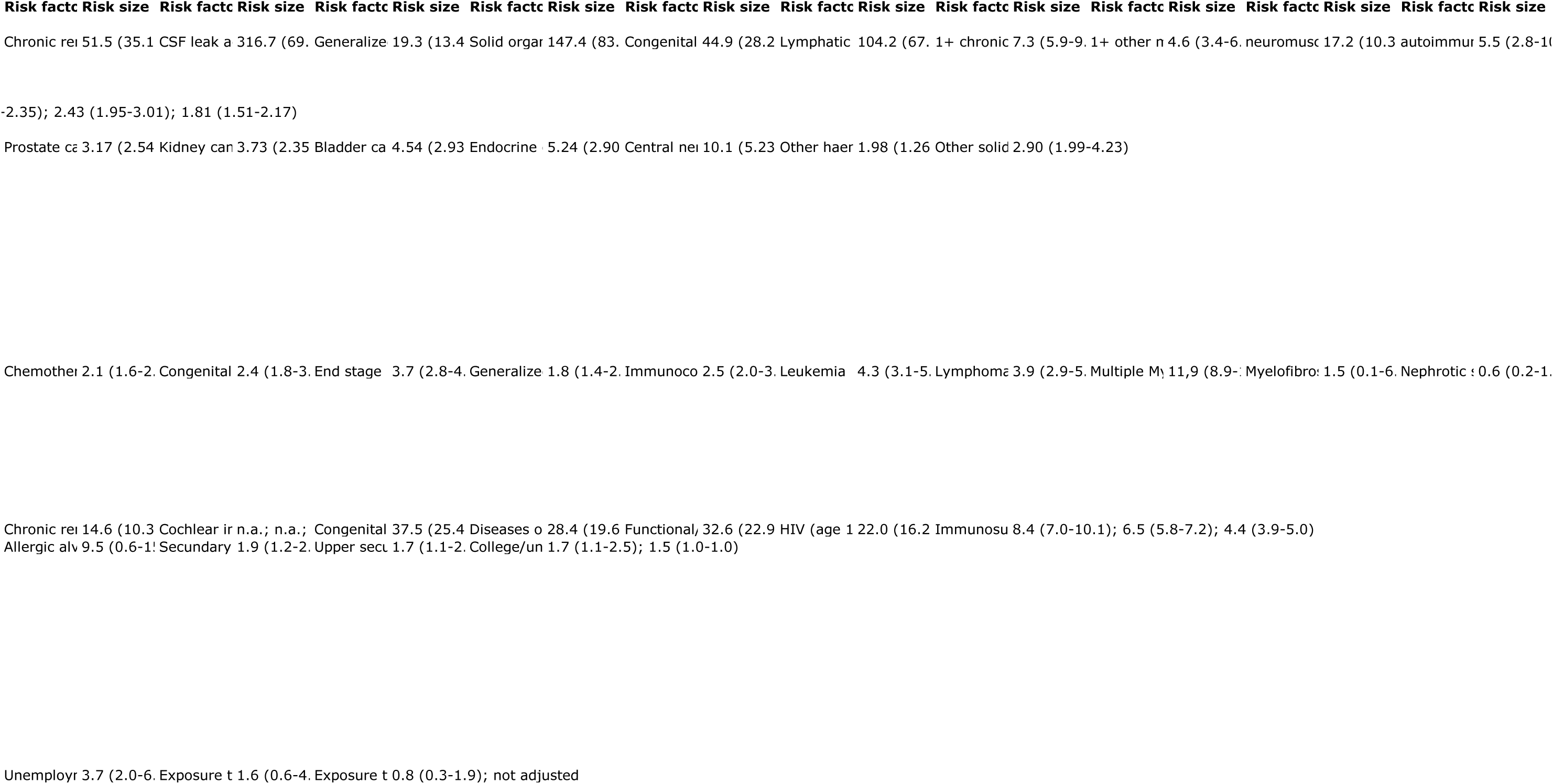
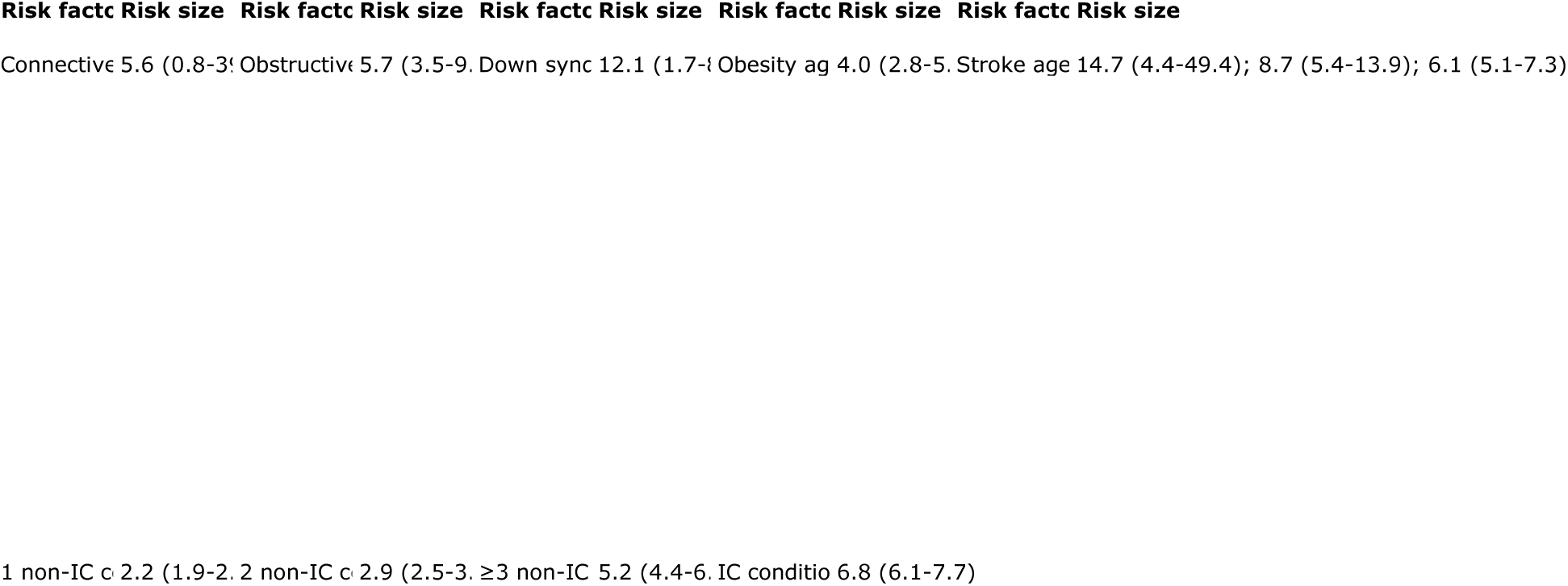

